# ClinSeg: Robust Brain Segmentation for Clinically Acquired Pediatric MRI

**DOI:** 10.64898/2026.08.28.26361643

**Authors:** Elizabeth Levitis, Henry F. J. Tregidgo, Dabriel Zimmerman, Benjamin Jung, Shivaram Karandikar, Margaret Gardner, Philip Mattisson, Eren Kafadar, Anna Zapaishchykova, Benjamin H. Kann, Susan T. Sotardi, Arastoo Vossough, Hao Huang, Benjamin Billot, Juan Eugenio Iglesias, Daniel C. Alexander, Aaron Alexander-Bloch, Jakob Seidlitz

**Affiliations:** Department of Child and Adolescent Psychiatry and Behavioral Sciences, Children’s Hospital of Philadelphia, Philadelphia, Pennsylvania; Neurodevelopment & Psychosis Section, Department of Psychiatry, Perelman School of Medicine, University of Pennsylvania, Philadelphia, Pennsylvania; Lifespan Brain Institute, Children’s Hospital of Philadelphia and Penn Medicine, Philadelphia, Pennsylvania; UCL Hawkes Institute, Department of Computer Science, University College London, London, United Kingdom; Artificial Intelligence in Medicine (AIM) Program, Mass General Brigham, Harvard Medical School, Boston, MA, USA; Department of Radiation Oncology, Dana-Farber Cancer Institute, Brigham and Women’s Hospital, Boston Children’s Hospital, Harvard Medical School, Boston, MA, USA; Department of Radiology, Children’s Hospital of Philadelphia, Philadelphia, Pennsylvania; Department of Radiology, Perelman School of Medicine, University of Pennsylvania, Philadelphia, PA, USA; Université Côte d’Azur, Inria, Epione team, Sophia Antipolis, France; Athinoula A. Martinos Center for Biomedical Imaging, Massachusetts General Hospital and Harvard Medical School, Boston, MA, United States; Computer Science and Artificial Intelligence Laboratory, Massachusetts Institute of Technology, Cambridge, MA, United States

## Abstract

Clinical brain MRIs from pediatric health systems represent a viable resource for modeling early neurodevelopmental trajectories and studying neurodevelopmental risk in real-world populations. However, a limitation to date has been the performance of existing segmentation tools for measuring various brain phenotypes in clinical scans. In particular, many tools underperform in infant scans due to morphological and physical changes such as rapid myelination. Here, we introduce ClinSeg: a robust segmentation approach tailored to early-life clinical MRIs with variable orientation, resolution, and contrast. We leverage existing registration and synthetic data generation tools to construct a training corpus for a 3d U-Net spanning anatomical and contrast diversity, including scans with morphological abnormalities from a pediatric hospital. Validated against manual segmentations, ClinSeg outperforms existing models in infancy while matching them in childhood and adolescence. Finally, ClinSeg enables the construction of reference brain growth trajectories in 11,699 individuals from 0-21 years of age, leading to the detection of more nuanced age-related findings in clinical groups.

## Introduction

Early infancy represents a critical period for brain development, encompassing rapid changes in myelination, folding, and synaptogenesis^1^. These processes are reflected in distinct growth trajectories observed in macrostructural phenotypes, such as cortical thickness, surface area, and grey matter volume^2^. Measurement of these phenotypes requires robust brain tissue segmentation of brain magnetic resonance images (MRIs) to delineate structural boundaries, and traditional segmentation algorithms typically fail when applied to early developmental data^3^.

Accurate segmentation of infant MRI scans necessitates overcoming challenges posed by the changing morphology of infant brains as well as the dynamic process of ongoing myelination, which begins in the fifth fetal month. Morphological changes include tertiary folding and rapid expansion of cortical surface area in the first year of life^4^. Many infants also undergo gross changes in head shape, such as the onset and resolution of positional plagiocephalies^5^. Simultaneously, myelination causes changes in the physical properties of gray and white matter tissue that manifest in three distinct patterns of gray and white matter contrast visible on T1-weighted (T1w) and T2-weighted (T2w) scans throughout the first year of life^6,7^. Adult gray-white tissue contrast is reversed in infancy, followed by a period of isointensity, and then the emergence of adult-like tissue contrasts. Moreover, there are significant inter-individual and inter-regional differences in the progression through these three patterns.

While most prior studies of infant brain segmentation have been constrained to prospective research protocols with high-quality, 1 mm isotropic acquisitions, applying these methods to heterogeneous clinically acquired MRIs remains a technical challenge. Compared with research-quality scans which tend to have high signal to noise ratios and uniform acquisition protocols, clinically acquired images exhibit substantial heterogeneity in sequence type, image contrast, and spatial resolution. Notably, SynthSeg and SynthSeg+ have addressed this challenge by leveraging convolutional neural networks (CNNs), specifically through the UNet architecture^8–10^, in conjunction with a contrast-agnostic approach for generating synthetic data, to enable segmentation of clinically acquired data of varying resolution and acquisition protocols^11^. In the pediatric context, we recently curated a large, retrospective clinical dataset from the Children’s Hospital of Philadelphia (CHOP) – the CHOP Scans With Limited Imaging Pathology (SLIP) cohort^12,13^. Deploying SynthSeg+ and related methods in CHOP SLIP accurately recapitulated childhood trajectories of brain development and deviations in clinical cases identified in large research-quality datasets^2,12–14^. However, unreliable segmentation resulted in the exclusion of most, if not all, individuals under 6 months of age^9,14^. Despite the potential of CHOP SLIP and other retrospective clinical datasets for modeling early neurodevelopment, the effective use of this data is constrained by the capability of current automated segmentation approaches to reliably segment a substantial proportion of infant brain MRIs.

While contrast-agnostic CNNs such as SynthSeg+ and UltimateSynth are robust to heterogeneous tissue contrasts, they lack infant-specific training data, which may be necessary to capture the range of morphological and myelination-related heterogeneity of infant brain MRI^8,9,15^. By contrast, the recently described Baby Infant Brain Segmentation (BIBSNet) model addresses this limitation by combining the contrast-agnostic synthetic data generation from the SynthSeg+ framework with additional training data from the Baby Open Brains (BOBs) dataset, the largest collection of infant MRI scans for individuals aged 0-8 months with accompanying manually corrected segmentations^16,17^. In a 10-fold cross-validation framework incorporating both real and synthetic scans, BIBSNet demonstrated improved segmentation performance relative to earlier methods such as Joint Label Fusion (JLF)^18^. However, BIBSNET was tested on synthetic data that is not representative of real-world clinical scans, with unrealistic simulations of contrasts, resolutions, heavy preprocessing, and limited age span.

Here, we address these gaps by developing ClinSeg, a robust segmentation approach explicitly tailored to early-life, clinically-acquired MRI, which also demonstrates strong performance in older individuals. We build upon the strengths of previous segmentation models such as SynthSeg+ and BIBSNet by integrating manually corrected labels from the BOBs dataset with the nnU-Net framework for optimized 3D U-Net training. We enhance the training data by leveraging a deep learning registration approach to map BOBs segmentations onto scans representing normative as well as morphologically extreme clinical data, and we employ ak-means augmentation strategy that simulates the evolving white matter myelination patterns characteristic of the first year of life. We validate ClinSeg across diverse research and clinical datasets, including clinical scans of infants with the 22q11.2 Deletion Syndrome (22q11DS), and demonstrate ClinSeg’s utility by constructing clinical neuroanatomical growth charts that cover the early infant period. Both segmentation models and growth charts are publicly shared (https://github.com/BGDlab upon publication) for use by other researchers.

## Results

We first demonstrate the efficacy of our segmentation model (Figure 1) on research-quality scans with accompanying manual segmentations, specifically showing robust performance in infant data and comparable performance in adult data. Next, we show that in scans with limited imaging pathology, ClinSeg’s performance surpasses that of state-of-the-art segmentation pipelines for individuals between 0-1 years of age and also successfully handles scans from older individuals. New growth charts are fit across global and regional phenotypes, demonstrating that we can simultaneously recapitulate previously reported trajectories while including infant data, enabling benchmarking of future scans.

**Figure 1.**
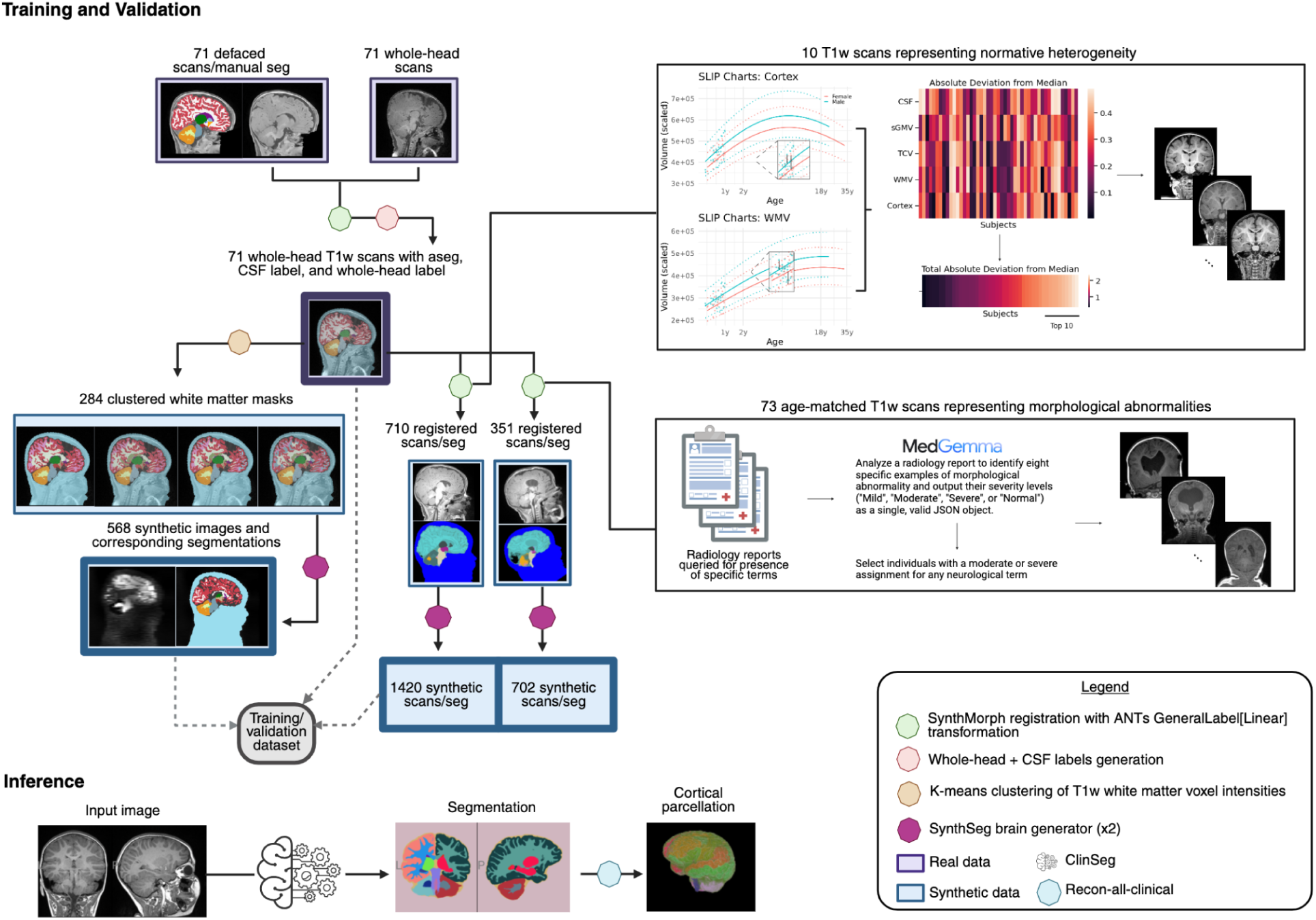
ClinSeg workflow for training/validation dataset curation and inference. We assemble a dataset of real and synthetic data for training a 3D UNet using the nnUNetv2 framework. Starting with 71 T1w scans and accompanying segmentations from the BOBs dataset, we register the scans back to native-space, transform the accompanying segmentations, and generate gross whole head labels. These updated scans/segmentations are used in three training data generation streams: 1) to generate scans representing normative heterogeneity, 2) to generate scans representing morphological abnormalities, 3) to generate scans encompassing heterogeneous white matter contrast. Cumulatively, the corpus of training data comprises 3822 scans and segmentations. The nnUNetv2 framework is used to train a 3D UNet with 5-fold cross validation. At inference time, the whole-head label is dropped, returning a volumetric segmentation. These segmentations can be plugged into the recon-all-clinical workflow of FreeSurfer to generate downstream surfaces and cortical parcellations.

### ClinSeg produces labels that closely resemble gold-standard manual segmentations

To assess the performance of the segmentation models, we benchmarked the models against expert manual segmentations of the Developmental Infant Brain Study (DIBS, 0-3 years, n=43) and the Child and Adolescent NeuroDevelopment Initiative (CANDI, 3-21 years, n=103).

Compared with BIBSNet and SynthSeg+, ClinSeg yielded improved or comparable segmentation of cerebral cortex and white matter in both datasets (Figure 2, Table S1). ClinSeg outperformed both BIBSNet and SynthSeg+ in the early developmental DIBS dataset, where manual segmentations were available for cerebral white matter and the cerebral cortex. Dice scores with manual segmentations were consistently higher for cerebral white matter (ClinSeg, mean = 0.909, sd = 0.0267; SynthSeg+, mean = 0.857, sd = 0.0132; BIBSNet, mean = 0.89, sd = 0.0367) and for cerebral cortex (ClinSeg, mean = 0.913, sd = 0.0269; SynthSeg+, mean= 0.86, sd = 0.014; BIBSNet, mean = 0.891, sd = 0.0363) (Figure 2a). The differences in Dice scores between the models were statistically significant based on a pairwise Friedman test (cerebral white matter: p = 1.23e-16, cerebral cortex: p = 4.22e-17) and reflected in observable improvements in segmentation quality upon visual inspection (Figure 2b). These results suggest high-quality performance of ClinSeg for the critical task of segmenting the cerebral gray-white boundary in early brain development.

**Figure 2.**
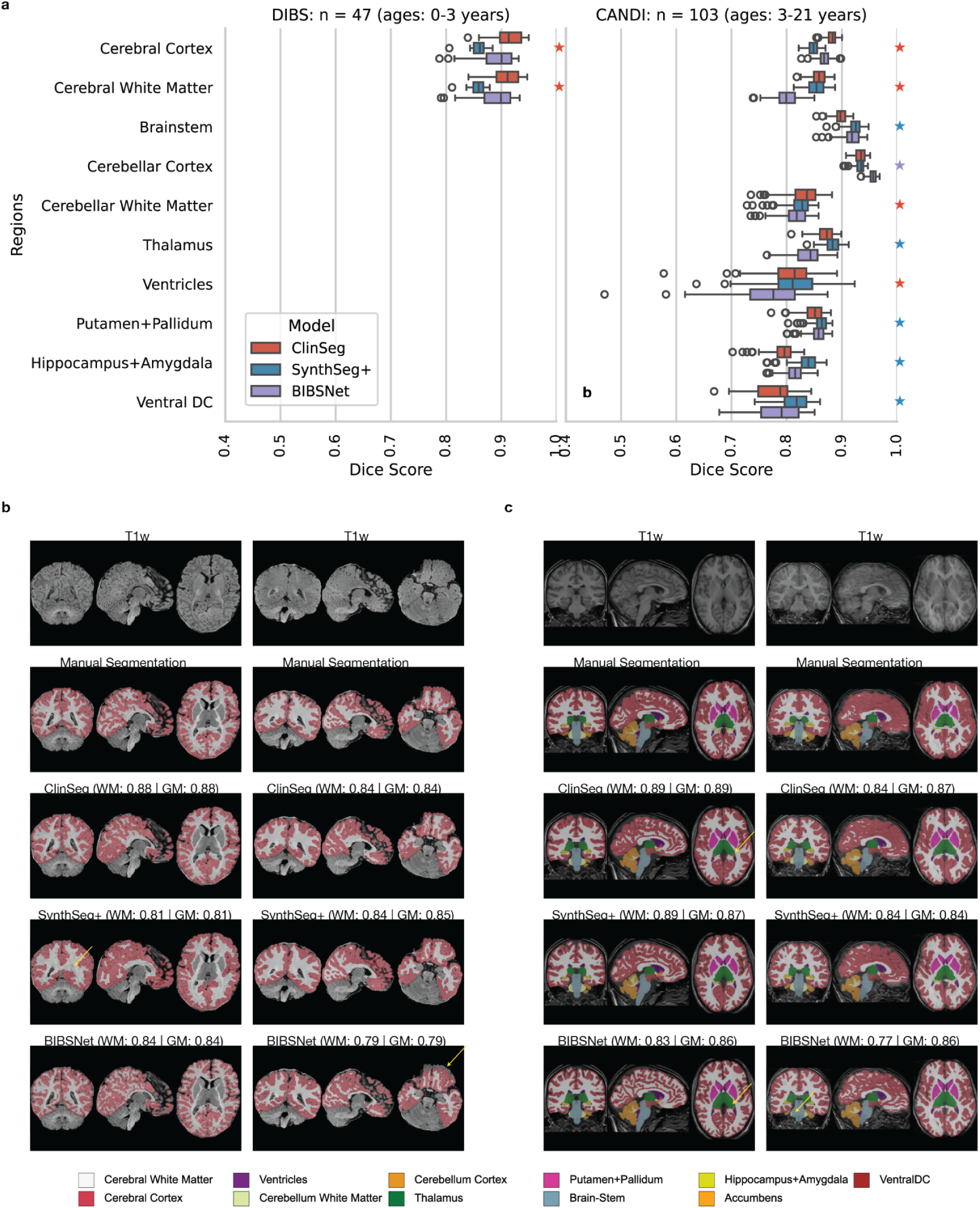
ClinSeg performance in datasets with manual segmentations. **(a)** Dice score performance comparing automated segmentation models to manual segmentation of two research quality datasets, an early infant dataset (DIBS) and a childhood/adolescent dataset (CANDI). Models include the present model (ClinSeg) as well as two widely used comparison models (SynthSeg+ and BIBSNet). To decrease the number of multiple comparisons, segmentations of the putamen and pallidum and segmentations of the hippocampus and amygdala were combined for the purposes of calculating the Dice scores. The color of the star per region signifies the best-performing model based on median dice score. **(b)** Example segmentations in the DIBS dataset. Yellow arrows point to incomplete cortex coverage by BIBSNet. Dice scores for composite cerebral grey and white matter are shown for the three automated segmentation models. **(c)** Example segmentations in the CANDI dataset. Panel on the left has a yellow arrow for ClinSeg and BIBSNet indicating where hippocampus and amygdala are labeled as cerebral grey matter (correctly segmented by SynthSeg+). Panel on the right has a yellow arrow pointing to oversegmentation of the brainstem by BIBSNet. Dice scores for composite cerebral grey and white matter are shown for the three automated segmentation models. Abbreviations: GM, gray matter; WM, white matter; Ventral DC, Ventral Diencephalon.

Although ClinSeg was trained for segmenting early developmental scans, we also assessed the model’s robustness for the task of segmenting scans in older age ranges. In the childhood/adolescent dataset CANDI, we observed similarly elevated performance for both ClinSeg (mean Dice = 0.86) and SynthSeg+ (mean Dice = 0.85) over BIBSNet (mean Dice = 0.8) for cerebral white matter (Fig 2a). More comparable performance was observed across pipelines for the cerebral cortex (mean Dice for ClinSeg, 0.88; mean Dice for BIBSNet, 0.87; mean Dice for SynthSeg+, 0.85). Dice performance for the subcortical and cerebellar structures was more variable, with lower performance for ClinSeg observed in the hippocampus and amygdala. Differences in Dice scores between the models were statistically significant for many regions (see Table S1) and reflected in observable differences in segmentation quality upon visual inspection (Figure 2c). Overall, these results suggest a high degree of robustness for ClinSeg for the task of segmenting scans in older age ranges, with comparable performance to existing models.

### ClinSeg sustains high segmentation and cortical parcellation performance in clinical data

Having demonstrated ClinSeg’s robust performance in research quality scans with manual segmentations, we subsequently evaluated it on a total of 78,508 clinical MRIs ages 0-21 from the Children’s Hospital of Philadelphia (CHOP) Scans with Limited Imaging Pathology (SLIP) cohort^13^. As described in the Methods, we first benchmark performance for segmentations produced by ClinSeg against those produced by SynthSeg+ implemented as part of the broader recon-all-clinical (RAC) tool shipped with the FreeSurfer suite^19^, which encompasses SynthDist^19^ for estimating surfaces from segmentations, along with FreeSurfer functionality for generating cortical parcellations. We ascertained robustness of both SynthSeg+ and ClinSeg to clinical acquisitions using the automated composite regional quality control (QC) scores described in Billot et al^9^, with a minimum threshold of 0.65 applied across nine regions used as the criteria for passing QC. Here, we report performance first based only on segmentation level QC.

ClinSeg yielded high-quality segmentations of early developmental clinical scans. Notably, no neonatal scans (<1 month of age) passed automatic QC using SynthSeg+, whereas 195 scans – comprising 98% of the total neonatal sample – passed QC using ClinSeg. ClinSeg also performed substantially better for infants aged 1-6 months, yielding an additional 2,523 passing scans across 556 subjects (564 sessions). Gains were also present for individuals aged 6–12 months, though less pronounced overall. We note that while the SynthSeg+ QC module was originally designed for adult clinical brain MRIs without prior validation for infant data, comparing QC scores against Dice scores for ClinSeg segmentations across the DIBS (ages 0-3) and CANDI (ages 3-21) datasets revealed consistent mapping (Figure S1). Specifically, in the cerebral cortex, the infant dataset (DIBS) demonstrated higher Dice scores [0.86, 0.95] corresponding to higher QC scores [0.72,0.78] compared to CANDI. Overall, strong Dice scores [0.82, 0.95] across cerebral cortex and white matter mapped to passing QC scores [0.65, 1.0]. Visual inspection confirmed that automated QC scores in SLIP indexed observable differences in segmentation quality throughout early development (Figure 3b; see Figure S2 for pass/fail examples across the first year of life).

**Figure 3.**
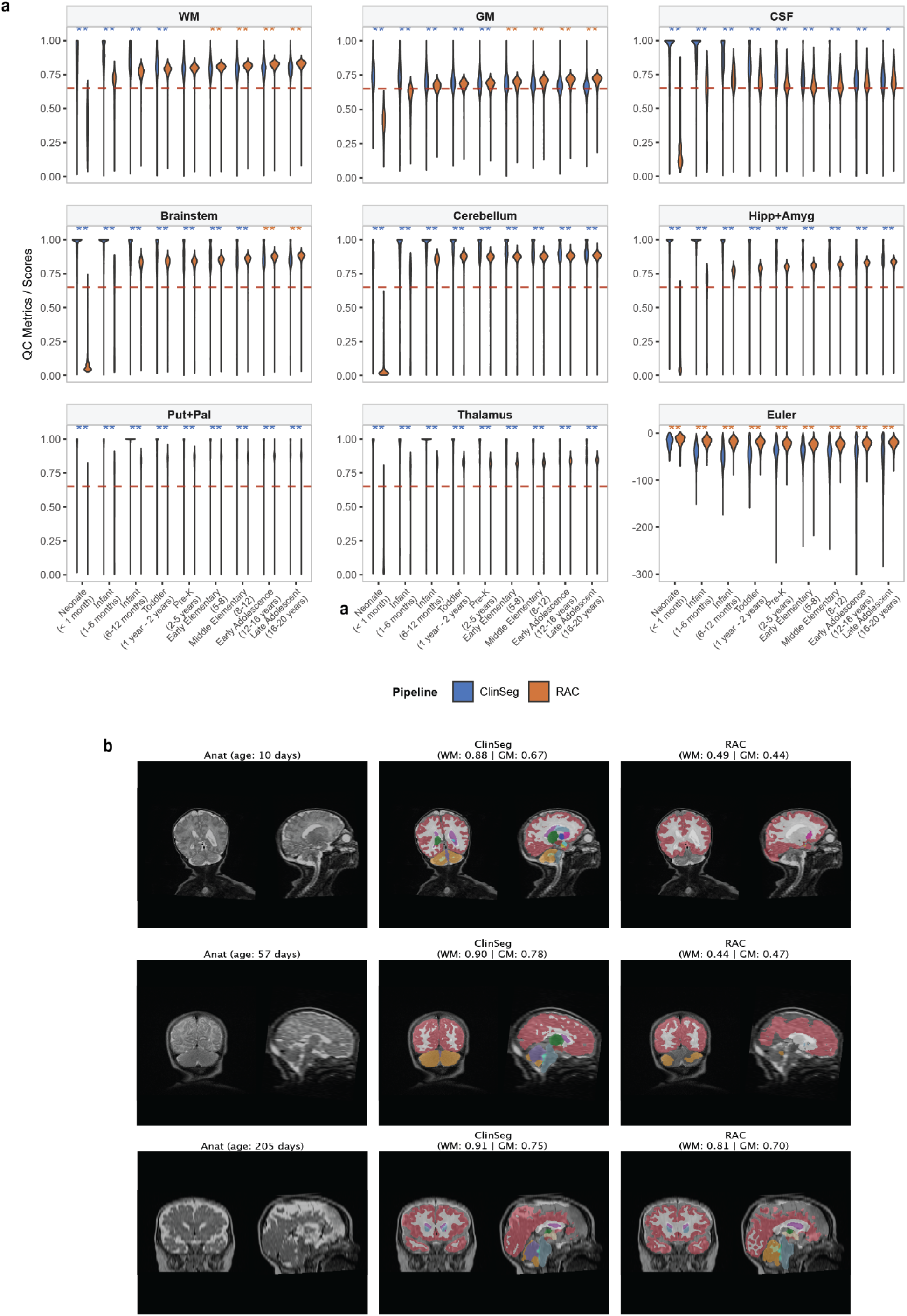
Performance of automated segmentation with ClinSeg versus SynthSeg+/RAC for clinical brain MRIs in the Children’s Hospital of Philadelphia Scans with Limited Imaging Pathology Cohort. **(a)** Distribution of predicted quality control (QC) scores for nine composite regions across eight age groups using either SynthSeg+ (orange) or ClinSeg (blue) segmentations. For all measures except for “Euler mean”, the dashed line corresponds to the recommended QC “pass” threshold of 0.65. **(b)** Example anatomical scan, ClinSeg segmentation, and SynthSeg+/RAC segmentation for a 22-day-old (neonate < 1 month category), 77-day-old (infant 1-6 month category), and 291-day-old (infant 6-12 month category). For the 22 and 77-day-old subjects, RAC fails to segment the ventricles, brainstem, and cerebellum. Captions include the predicted QC scores for white matter (WM) and grey matter (GM) from the SynthSeg+ QC module.

ClinSeg also showed a high degree of robustness in segmenting MRIs in older age ranges. Although more individuals passed QC with SynthSeg+ in the 12-21 age range, the overall difference of 157 individuals was small relative to the number of individuals in this age range in the cohort. The performance discrepancy between the two pipelines varies systematically with age and acquisition type (Fig 3a, Fig S1). SynthSeg+ begins outperforming ClinSeg on predicted grey matter segmentation quality by age 5 for non-MPRAGE T1w and T2w scans, but not until age 12 for MPRAGE and FLAIR scans. For predicted cerebrospinal fluid segmentation, SynthSeg+ only outperforms ClinSeg for MPRAGE scans in the oldest age category of 16-21 years.

As seen in Table 1, total scans passing segmentation and Euler QC were lower for ClinSeg (42,953) compared with SynthSeg+ (48,230); however, ClinSeg enabled a total of 952 additional subjects’ data to be retained.

**Table 1.** Increased number of clinical scans pass quality control (QC) for automated segmentation with ClinSeg compared to recon-all clinical (RAC). Aggregate scans, participants, and sessions passingQC for ClinSeg and RAC. We provide total scan, participant, and session counts for all data processed, data passing segmentation QC only, and data passing segmentation QC (seg QC, based on synthseg’s quality control module) and Euler number based QC (based on surface holes in the cortical reconstruction).

|  | Scans<sup>1</sup> |  |  | Sessions |  |  | Participants |  |  |
| --- | --- | --- | --- | --- | --- | --- | --- | --- | --- |
|  | RAC | ClinSeg | Δ | RAC | ClinSeg | Δ | RAC | ClinSeg | Δ |
| **Processed** |  |  |  |  |  |  |  |  |  |
| Neonate (< 1 month) | 640 | 1,226 | 586 | 193 | 200 | 7 | 193 | 200 | 7 |
| Infant (1-6 months) | 2,809 | 3,803 | 994 | 558 | 580 | 22 | 552 | 571 | 19 |
| Infant (6-12 months) | 3,316 | 3,746 | 430 | 568 | 592 | 24 | 565 | 588 | 23 |
| Toddler (1-2 years) | 5,231 | 5,678 | 447 | 863 | 876 | 13 | 854 | 864 | 10 |
| Pre-K (2-5 years) | 8,758 | 9,449 | 691 | 1,450 | 1,471 | 21 | 1,393 | 1,411 | 18 |
| Early Elementary (5-8) | 8,383 | 9,053 | 670 | 1,402 | 1,422 | 20 | 1,375 | 1,391 | 16 |
| Middle Elementary (8-12) | 11,657 | 12,537 | 880 | 1,964 | 1,975 | 11 | 1,908 | 1,919 | 11 |
| Early Adolescence (12-16 years) | 18,322 | 19,129 | 807 | 3,485 | 3,500 | 15 | 3,299 | 3,314 | 15 |
| Late Adolescent/Early Adult (16-21 years) | 13,286 | 13,887 | 601 | 2,551 | 2,564 | 13 | 2,417 | 2,430 | 13 |
| Total | 72,402 | 78,508 | 6,106 | 13,034 | 13,180 | 146 | 12,556 | 12,688 | 132 |
| **Seg QC only** |  |  |  |  |  |  |  |  |  |
| Neonate (< 1 month) | 0 | 904 | 904 | 0 | 195 | 195 | 0 | 195 | 195 |
| Infant (1-6 months) | 562 | 3,085 | 2,523 | 181 | 564 | 383 | 180 | 556 | 376 |
| Infant (6-12 months) | 2,085 | 2,795 | 710 | 500 | 558 | 58 | 499 | 556 | 57 |
| Toddler (1-2 years) | 3,690 | 4,192 | 502 | 810 | 856 | 46 | 803 | 847 | 44 |
| Pre-K (2-5 years) | 5,428 | 6,098 | 670 | 1,300 | 1,427 | 127 | 1,249 | 1,372 | 123 |
| Early Elementary (5-8) | 4,832 | 5,442 | 610 | 1,221 | 1,374 | 153 | 1,199 | 1,349 | 150 |
| Middle Elementary (8-12) | 7,078 | 7,069 | -9 | 1,737 | 1,905 | 168 | 1,688 | 1,852 | 164 |
| Early Adolescence (12-16 years) | 14,011 | 10,572 | -3,439 | 3,322 | 3,264 | -58 | 3,147 | 3,105 | -42 |
| Late Adolescent/Early Adult (16-21 years) | 10,772 | 7,178 | -3,594 | 2,459 | 2,325 | -134 | 2,332 | 2,217 | -115 |
| Total | 48,458 | 47,335 | -1,123 | 11,530 | 12,468 | 938 | 11,097 | 12,049 | 952 |
| **Seg QC + Euler<sup>2</sup>** |  |  |  |  |  |  |  |  |  |
| Neonate (< 1 month) | 0 | 275 | 275 | 0 | 121 | 121 | 0 | 121 | 121 |
| Infant (1-6 months) | 562 | 2,258 | 1,696 | 181 | 523 | 342 | 180 | 516 | 336 |
| Infant (6-12 months) | 2,082 | 2,568 | 486 | 500 | 552 | 52 | 499 | 550 | 51 |
| Toddler (1-2 years) | 3,667 | 3,891 | 224 | 810 | 848 | 38 | 803 | 840 | 37 |
| Pre-K (2-5 years) | 5,384 | 5,875 | 491 | 1,299 | 1,416 | 117 | 1,248 | 1,361 | 113 |
| Early Elementary (5-8) | 4,800 | 5,245 | 445 | 1,217 | 1,366 | 149 | 1,195 | 1,341 | 146 |
| Middle Elementary (8-12) | 7,039 | 6,860 | -179 | 1,737 | 1,890 | 153 | 1,688 | 1,839 | 151 |
| Early Adolescence (12-16 years) | 13,947 | 9,679 | -4,268 | 3,322 | 3,227 | -95 | 3,147 | 3,071 | -76 |
| Late Adolescent/Early Adult (16-21 years) | 10,749 | 6,302 | -4,447 | 2,459 | 2,285 | -174 | 2,332 | 2,182 | -150 |
| Total | 48,230 | 42,953 | -5,277 | 11,525 | 12,228 | 703 | 11,092 | 11,821 | 729 |
| **Seg QC + Euler + GAMLSS** |  |  |  |  |  |  |  |  |  |
| Neonate (< 1 month) | 0 | 262 | 262 | 0 | 117 | 117 | 0 | 117 | 117 |
| Infant (1-6 months) | 551 | 2,192 | 1,641 | 179 | 513 | 334 | 178 | 506 | 328 |
| Infant (6-12 months) | 2,081 | 2,472 | 391 | 500 | 551 | 51 | 499 | 549 | 50 |
| Toddler (1-2 years) | 3,641 | 3,642 | 1 | 805 | 839 | 34 | 798 | 831 | 33 |
| Pre-K (2-5 years) | 5,320 | 5,440 | 120 | 1,285 | 1,399 | 114 | 1,236 | 1,346 | 110 |
| Early Elementary (5-8) | 4,780 | 4,963 | 183 | 1,213 | 1,346 | 133 | 1,191 | 1,321 | 130 |
| Middle Elementary (8-12) | 6,985 | 6,571 | -414 | 1,724 | 1,871 | 147 | 1,675 | 1,820 | 145 |
| Early Adolescence (12-16 years) | 13,854 | 9,393 | -4,461 | 3,301 | 3,197 | -104 | 3,131 | 3,046 | -85 |
| Late Adolescent/Early Adult (16-21 years) | 10,712 | 6,158 | -4,554 | 2,450 | 2,266 | -184 | 2,323 | 2,163 | -160 |
| Total | 47,924 | 41,093 | -6,831 | 11,457 | 12,099 | 642 | 11,031 | 11,699 | 668 |

<sup>1</sup> Δ = ClinSeg - RAC. Blue = ClinSeg retains more; red = RAC retains more.
<sup>2</sup> Seg QC threshold: > 0.65. Euler QC: modified z-score > -3.5.

### ClinSeg enables the new creation of clinical brain growth charts of infancy

Ultimately, an overarching goal for leveraging clinically acquired scans is to benchmark global and regional brain phenotypes against a clinical reference. We sought to improve upon previous versions of SLIP-based reference charts by leveraging ClinSeg’s success in segmenting infant data to expand the age range of our growth charts^12,14^. Beyond looking at gross tissue changes in the cerebrum, we sought to get cortical parcellations and cortical phenotypes beyond volume (e.g. thickness and surface area) to enable comparison with past work^2,12,14^. As elaborated on in the Methods, recon-all-clinical natively uses segmentations generated by SynthSeg+ as an input to its surface reconstruction module, and we adapt this workflow using segmentations from ClinSeg^9,19^. We used SLIP scans that passed QC for both the aforementioned segmentation-based quality control measures, the surface-based Euler number, and had nonzero values for regional volumes (see Table 1, Seg QC + Euler + GAMLSS filtering for scan counts). Next, we modeled global and regional phenotypes using generalized additive models for location, scale, and shape (GAMLSS) with penalized splines and a Box-Cox t distribution (see Methods)^20^. As a point of comparison, we repeated this modeling using SynthSeg+/RAC (Table 1 for the sample size; Figure S5). Here, we describe outputs for both global and regional features following the RAC-informed cortical refinement and present results for global features directly from ClinSeg in the supplementary materials (Figure S6).

Across the global measures we considered – specifically, cerebral tissue compartments such as grey matter, white matter, subcortical grey matter, and ventricular volumes – worm plots indicated a better fit for ClinSeg compared with RAC (Figure S4). Additionally, milestones were largely consistent with those reported for models fit on a global, lifespan sample of research-quality scans (Figure 4a-d; Supplemental Table 2). Regional cortical peak maturations recapitulated the previously reported developmental gradient across the cortex. The earliest maturing structure was identified as the left transverse temporal region which is part of the primary auditory cortex, peaking at 3.97 years of age. Other early-peaking structures (maturing prior to age 6) were predominantly localized to primary sensory and motor regions. These included the bilateral lingual gyrus (left hemisphere (LH): 4.26, right hemisphere (RH): 5.59) and bilateral precuneus (LH: 5.77, RH: 5.86). In contrast, delayed peak maturations were concentrated largely within higher-order association areas, with the exception of precentral gyrus, peaking between 9 and 13 years of age. These late-maturing regions included the caudal and rostral anterior cingulate cortices, insula, caudal middle frontal cortex, entorhinal cortex, and the parahippocampal cortex, the latter of which represented the latest overall maturing region with a peak at 13.3 years of age (Figure 4e-g). We noted strong agreement for centiles estimated across regional volumes between ClinSeg and RAC (mean ICC = 0.82, sd = 0.11; Figure S7).

**Figure 4.**
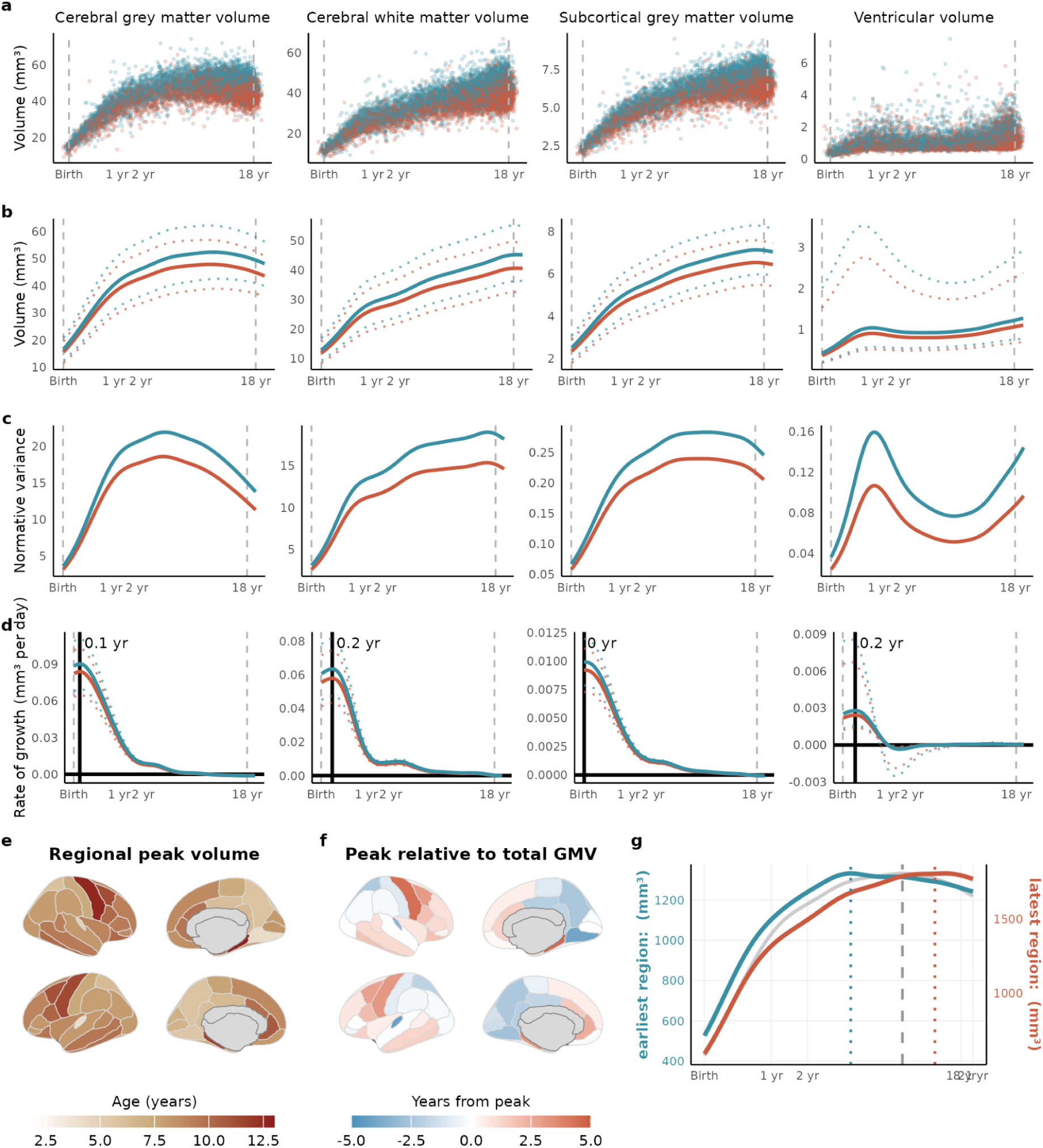
Clinical brain MRI growth charts for ClinSeg-derived global tissue classes. **(a)** Raw bilateral cerebrum tissue volumes for grey matter, white matter, subcortical grey matter and ventricles are plotted for each session’s data as a function of age (log-scaled) with points colored by sex (female, red; male, blue). **(b)** Normative brain volume trajectories were estimated using GAMLSS modeling as described in Methods. The dotted trajectories represent the 2.5% and 97.5% centiles. **(c)** Trajectories of median between-subject variability are estimated as the predicted dispersion from GAMLSS. **(d)** Rates of volumetric change for each sex-specific trajectory were estimated using the first derivative of the median volumetric trajectory. Solid horizontal lines denote the tissue-specific volume at which the tissue begins to shrink while the solid vertical line denotes the age of maximum growth. **(e)** Regional peak volumes for the 68 Desikan-Killiay cortical regions. **(f)** Regional peaks relative to GMV peak, where GMV peak is subtracted from each regional peak. **(g)** Trajectories for the earliest peaking region (left transverse temporal blue line) and the latest peaking region (right parahippocampal, red line), showing the range of regional variability relative to the GMV trajectory (grey line). Regional volume peaks are denoted as dotted vertical lines either side of the global peak, denoted as a dashed vertical line, in the bottom panel. The left *y* axis on the bottom panel refers to the earliest peak (blue line); the right *y* axis refers to the latest peak (red line). The y-axes in **a-d** are scaled in units of 10,000 mm^3^ (10 ml).

### Infant brain growth in 22q11.2 Deletion Syndrome

To further evaluate the impact of ClinSeg and its resulting growth charts, we applied them to 22q11.2 Deletion Syndrome (22q11DS) – a condition extensively studied from childhood through adulthood, but where infant data is missing from previous studies. Using ClinSeg, we repeated a case-control analysis using age- and sex-matched SLIP controls to evaluate structural brain alterations, previously performed in Jung et al. using RAC^14^. Of the 92 subjects that had previously passed QC under RAC, 83 also passed under ClinSeg; ClinSeg additionally allowed for the inclusion of 31 new subjects – including 8 aged 0–6 months and 7 aged 6 months–1 year (mean age = 1.98 years, SD = 2.84) – enabling one of the first investigations of early brain development in 22q11DS (Fig 5a). Demographics for both the previously included and newly added participants are in Table 2.

**Table 2.** Demographics for 22q11DS case-control analysis. The sample size and associated ages are reported for the current case-control sample included using ClinSeg as well as the sample of subjects overlapping with the sample previously included in Jung et al, 2026 (using the RAC pipeline).

| Sample Sizes and Age Distributions |  |  |  |  |
| --- | --- | --- | --- | --- |
| By segmentation method and diagnostic group |  |  |  |  |
|  | N |  | Mean Age [Min–Max]<sup>1</sup> |  |
|  | Control | 22q11DS | Control | 22q11DS |
| **Median** |  |  |  |  |
| ClinSeg | 400 | 114 | 5.74 [0.00–19.72] | 6.07 [0.00–20.33] |
| Overlap with RAC \| Jung et al, 2026<sup>2</sup> | 218 | 83 | 8.63 [0.78–19.62] | 7.78 [0.50–20.33] |
| **MPR** |  |  |  |  |
| ClinSeg | 191 | 56 | 6.72 [0.01–20.37] | 6.95 [0.01–20.33] |
| Overlap with RAC \| Jung et al, 2026<sup>2</sup> | 92 | 44 | 8.23 [1.10–19.18] | 7.95 [0.50–20.33] |

<sup>1</sup> Age reported as mean [min–max], in years.
<sup>2</sup> "Overlap with sample included in Jung et al, 2026.

**Figure 5.**
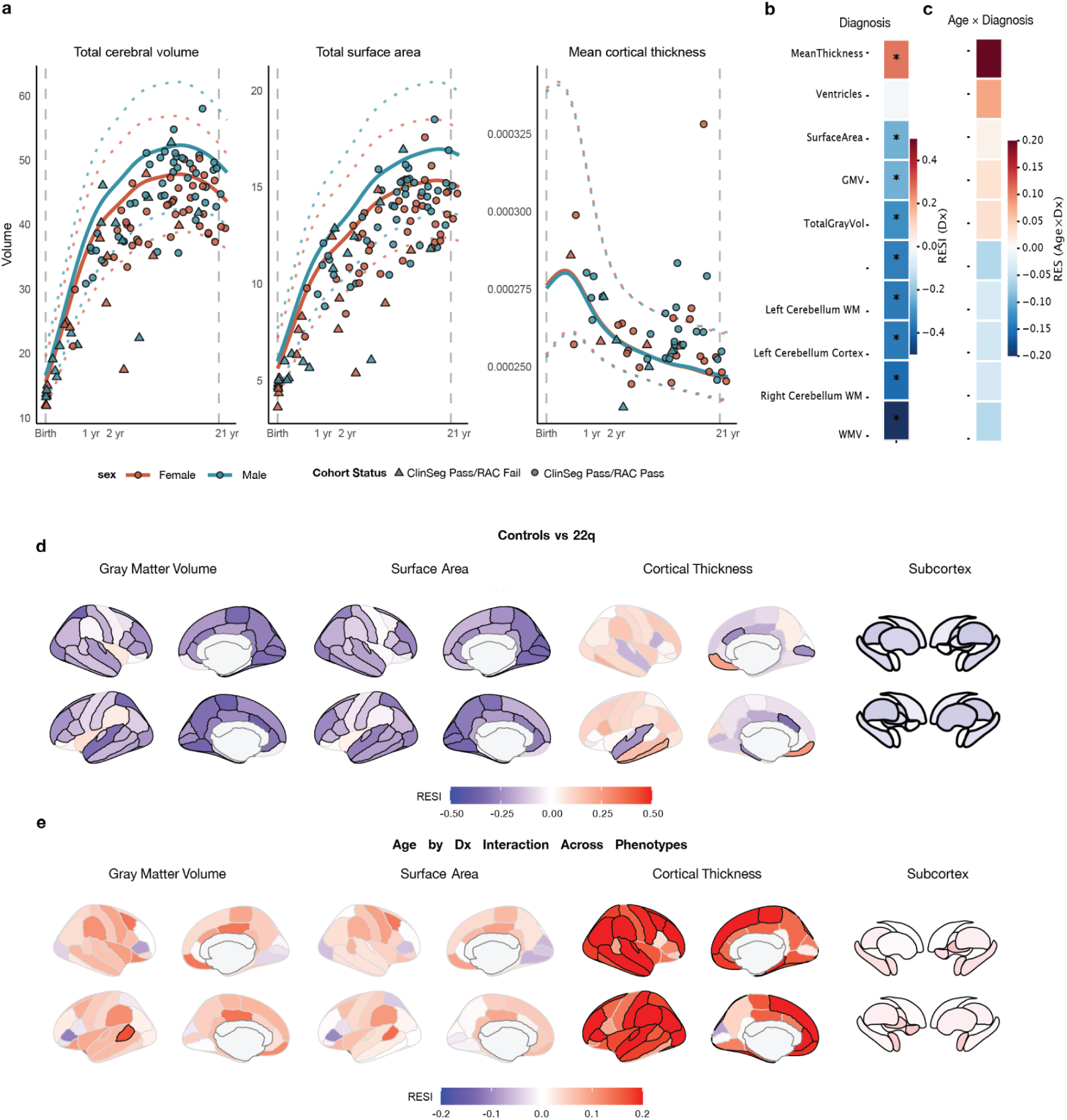
Segmentation performance and growth charts in the CHOP scans with limited imaging pathology (SLIP) cohort. (a) Normalized phenotype values plotted for 22q11DS cases over the SLIP derived growth curves. Individuals whose data passes QC for ClinSeg but not RAC are denoted by triangles, and those who pass QC for both are denoted by circles. (b) Robust effect size indices (RESI) are displayed for both case-control effects as well as (c) age by diagnosis interaction effects across the global and cerebellum measurements (d) RESI effect sizes are displayed for case-control effects for cortical grey matter volume, surface area, cortical thickness, and subcortical volumes. (e) age by diagnosis interaction effects for the same measurements. Bold outlines for cortical and subcortical structures indicate statistical significance after FDR correction (p_FDR_ < 0.05).

For metrics derived from the global measurements (*N_cases_*=114, *N_controls_*=400), 22q11DS was associated with significantly reduced total gray matter, subcortical gray matter, white matter volume, and surface area (all *p_FDR_* < 10^-3^), replicating prior results in Jung et al. Conversely, mean cortical thickness (CTh), isolated to the MPRAGE-only sub-cohort (N_cases_=56, N_controls_=191), was significantly higher in patients (Robust Effect Size Index (RESI) = 0.26, *p_FDR_* = 10^-3^; Figure 5b)^21^. In contrast to prior work, ventricular volumes showed no significant case-control differences (Figure 5b). There were no significant age-by-diagnosis interaction effects for the global measurements (Figure 5c).

Overall, regional effect sizes were highly concordant with those reported in Jung et al (GMV, r = 0.90; SA, f = 0.92; CTh, r = 0.90; Figure S8). Cortical GMV and SA deviations were decreased across most of the cortex and largely followed a convergent caudal to rostral gradient as previously reported. MPRAGE and multi-modality median-based models were highly consistent for GMV (r = 0.88) and SA (r = 0.89; Figure S9). For SA, the largest effect sizes seen in occipital regions such as the bilateral cuneus, lingual and pericalcarine areas (RESI = [-0.33, −0.36], *p_FDR_* < 10^-9^) and smallest in frontal regions such as the right pars opercularis, banks of the superior temporal sulcus, and medial orbitofrontal areas (RESI = [-0.109,-0.122], *p_FDR_* < 0.05; Figure 5d). 22q11DS showed greater reductions in GMV in two parietal areas (left paracentral region, RESI = −0.36, *p_FDR_* < 10^-11^; right superior parietal region, RESI = −0.36, *p_FDR_* < 10^-10^) as well as the left caudal anterior cingulate (RESI = −0.37, *p_FDR_* = 10^-11^). Additionally, while higher insula volumes persisted for SA and GMV, these deviations were not significant as reported previously. For GMV, no regions showed significant age by diagnosis interactions. For SA, we found significant age-by-diagnosis interaction effects only for the right parahippocampal region (RESI = −0.25, *p_FDR_* = 0.02) and the left superior parietal region (RESI = −0.22, *p_FDR_* = 0.048).

For CTh, case-control deviations were more variable, with more regions showing negative effect sizes than found in our previous work. In particular, the most divergent effect sizes for CTh were seen in the left caudal anterior cingulate (RESI = −0.306, *p_FDR_* < 10^-7^) while the left rostral middle frontal region showed increased CTh (RESI = 0.17, *p_FDR_* = 0.0037). Interestingly, we found significant and positive age-by-diagnosis interaction effects across much of the cortex for cortical thickness (RESI: [0.15, 0.26]), with little divergence between controls and individuals with 22q11DS early on followed by an increased divergence with development (Fig 5e; Figure S10). Given that our growth chart-derived centiles are measuring deviation from normality, this suggests that the higher thickness documented in individuals with 22q11DS emerges after infancy.

## Discussion

We present a segmentation model, ClinSeg, that enables volumetric analysis of clinical brain MRI scans in infancy. ClinSeg also performs well on scans through childhood, adolescence, and early adulthood, allowing a single segmentation model to be used throughout the pediatric period. We provide reference charts for ClinSeg-derived phenotypes, which recapitulate known global and regional volumetric trajectories from large research consortia and enable precise benchmarking of individual scans. Applying these tools to clinical brain MRIs of youth with 22q11.2 deletion syndrome (22q11DS), ClinSeg enables a large number of infant scans to be included in a case-control analysis that reproduces previously reported case-control differences and reveals novel age-by-diagnosis interactions.

The improved performance of ClinSeg in infant scans compared to other segmentation models is a function of its multipronged training data. ClinSeg adapts training based on synthetic brain MRIs generated from gold-standard segmentations, introduced by Synthseg, to early brain development. In early life, myelination increases in a spatially non-uniform manner, with different tracts undergoing myelination at different rates. By introducing distinct clusters of white matter labels within the white matter compartment, we generated a corpus of synthetic scans with more heterogeneous white matter contrast compared to other infant brain segmentation models, using a similar approach that was shown to be successful for a fetal adaption of SynthSeg^22,23^.

In addition to heterogeneous white matter contrast, ClinSeg’s training regimen included a wider range of brain anatomical variation, to mimic real-world variability in brain anatomy observed in clinical MRIs of infants. By deploying SynthMorph to nonlinearly register scans with gold standard manual segmentations to real clinical scans obtained at CHOP, with and without categorical abnormalities observed by pediatric neuroradiologists, ClinSeg’s training corpus included “real” looking scans with more heterogeneous morphometry compared to alternative segmentation models^9,23,24^. By exposing ClinSeg to more abnormal-looking morphology, for instance, we forced the model to learn how to handle greater variation in ventricle size as well as variation in sulcal width. Thus, ClinSeg attains superior performance on both research-quality and clinical-quality infant brain MRI scans.

Complementing its performance on infant scans, ClinSeg also performs well on brain MRIs obtained during early and late childhood, adolescence, and young adulthood. Most prior work that spans the pediatric age range relies on different segmentation models for scans in infancy compared to later developmental epochs, which potentially confounds effects of age and model performance^2,12^. To our knowledge, our ClinSeg brain charts are the first which span the entire pediatric age range using segmentations from a single model. Nonetheless, SynthSeg+ does achieve higher automated quality-control estimates for cerebral grey and white matter on scans from older participants, particularly for anisotropic T1-weighted and T2-weighted sequences.

This likely reflects ClinSeg having been exposed preferentially to training data constructed from infant segmentations. Constructing a training dataset from manual segmentations spanning a wider age range would be expected to improve performance on older scans, though such an approach might also degrade performance on infant scans.

Although ClinSeg’s core application is volumetric segmentation, we also use its output to initialize SynthDist, the surface-reconstruction component of the recon-all-pipeline, roducing extended cortical phenotypes from the ClinSeg segmentation^19^. SynthDist uses a deep learning prediction of the distance between white matter and pial surface combined with geometric processing to ensure a topologically accurate reconstruction of the cortical surface. This functionality enables surface-based cortical parcellation as well as phenotyping of cortical surface area (SA) and cortical thickness (CT). SynthDist was validated primarily on adult neuroanatomy, and its default pairing with SynthSeg+ segmentations (as part of recon-all-clinical) performs poorly on our SLIP cohort. This is likely a result of SynthSeg+’s white matter segmentations not being accurate enough to reliably initialize the white matter surface. Substituting ClinSeg’s more accurate white matter segmentation for this initialization improves the robustness of the resulting surface reconstruction.

Ultimately, ClinSeg allows for the inclusion of clinical infant brain MRIs in quantitative neuroimaging analyses, a major advantage for studies of early brain development as illustrated by our analysis of 22q11DS. By applying ClinSeg to a sample of clinical scans obtained at CHOP, we replicate past findings of reduced SA and grey matter volume (GMV) and find more nuanced alterations in CT than previously reported in 22q11DS. A recent study in an overlapping sample used Synthseg+ segmentations, necessitating the removal of infant scans due to poor quality segmentations. In contrast to this prior study, we find evidence of age-by-diagnosis interaction for CT, such that 22q11DS’s canonical thicker-than-normal cortex only emerges after infancy in early childhood, a finding that may reflect differential myelination, neuropil formation or synaptic pruning. Because our sample is cross-sectional, this interaction should be interpreted with caution, as we do not directly show within-individual differences in CT trajectories and find more nuanced group-level alterations in CT than previously reported in 22q11DS^25,26^.

Several limitations warrant future improvements to the ClinSeg model. Firstly, the quality of the segmentations depends heavily on the training data, and there remains a paucity of gold standard manual segmentations of early developmental brain MRIs, especially for infants with abnormal neuroanatomy. While we address this challenge using synthetically generated brain MRIs and registration of training data to real-world clinical scans with and without anatomical abnormalities, a larger corpus of high-quality manual segmentations may result in improvements to future segmentation models. Secondly, we adapted SynthDist and the automated QC module from Synthseg+ to ClinSeg, but both of these modules were originally trained on adult-like synthetic data. While the suboptimal automated QC measures may be biasing the pass rate of ClinSeg, visual evaluation of segmentations and corresponding QC scores demonstrate the qualitatively improved performance of ClinSeg in younger individuals (Figure S2). It is possible that specifically training pediatric versions of these modules would result in improved performance. Finally, ClinSeg is trained using the self-configuring nnUNet architecture. Adapting neuroimaging foundation models may yield superior segmentations in future work^27^.

Notwithstanding these limitations, ClinSeg represents a significant advance in segmenting early developmental clinical brain MRIs. There is now enormous potential to repurpose large clinical archives of infant brain MRIs for secondary research, providing a complementary methodological framework to large prospective research studies of early brain development^28^. ClinSeg’s segmentation model and the resulting growth charts are publicly shared for use by other researchers (upon publication).

## Methods and Materials

### Segmentation Model Conceptualization and Training

As described in the Introduction, substantial improvements in the space of segmentation for clinically acquired scans have come about with the advent of SynthSeg+ and its accompanying brain generator which enables the generation of synthetic data from existing segmentations using a domain-agnostic framework^9^. BIBSNet further extended this by training on infant data, and the SynthMorph deep-learning based registration framework opens up an opportunity to include examples of both normative heterogeneity and abnormal morphology^24^. In addition, the nnUNet framework has become a popular choice for training 3D UNet models by automatically preprocessing training data and choosing hyperparameters for model training^10,29^. In the next sections, we go into more detail about how the training data for ClinSeg was assembled.

### Training Datasets

The Baby Open Brains (BOBs) dataset comprised 71 defaced T1-weighted and T2-weighted scans from the Baby Connectome Project (BCP) with corresponding manual segmentations from infants aged 1-8 months^17,23^. These segmentations were re-registered to the corresponding non-defaced T1-weighted and T2-weighted scans from BCP to provide training data in native space. This dataset provided ground truth segmentations for building our training data.

To increase robustness to morphological abnormalities that may be enriched in clinical datasets, we first selected 10 subjects aged 0-1 years from the CHOP Scans with Limited Imaging Pathology (SLIP) cohort, hereafter referred to as SLIP-train^12,13^. These patients with clinically acquired T1-weighted scans were chosen based on specific criteria: scans had been segmented using SynthSeg+, passed SynthSeg+ quality control (minimum pseudo-Dice score of 0.65 across nine brain regions), and demonstrated elevated composite centile scores based on absolute deviation from the median (50th percentile) across cerebrospinal fluid (CSF), total cerebral volume (TCV), subcortical gray matter volume (sGMV), and white matter volume (WMV)^9^.

We additionally identified a set of CHOP patients (0-1 years) with categorical morphological abnormalities, hereafter referred to as CHOP-path, through automated review of radiology reports. Initially, we used SQL queries to search the narrative and impression text of radiology reports for specific terms including "benign enlargement of the subarachnoid space”, “Chiari malformation”, “ventriculomegaly”, and “sulcal prominence” (see Supplemental Materials for the full MedGemma prompt). This search identified 241 individuals with positive mentions of these terms in their reports. Subsequently, we employed a HIPAA-compliant instance of MedGemma accessed within the CHOP network to grade the severity of each condition mentioned in the reports, classifying each term as normal (absent), mild, moderate, or severe based on the radiological findings^30^. From this analysis, we identified 73 individuals who presented with severe manifestations of at least one categorical morphological abnormality. Figure 1 illustrates the process by which we used the BOBs scans in conjunction with the SLIP-train and CHOP-path scans to construct the training and validation dataset.

### Data Processing and Registration for Training Data

Segmented BOBs T1-weighted scans were registered back to their original non-defaced BCP space using SynthMorph to ensure whole-head coverage, and label maps were transferred from the BOBs to the BCP space using ANTs with label-preserving interpolation, producing BOBs-BCP pairs^24,31^. This registration process created the BOBs-BCP sample, establishing spatial correspondence between the manual segmentations and the BCP reference space. To avoid skull-stripping as a preprocessing step, we used a retrained SynthSeg+ model where training data had extracerebral labels segmented using the CHARM toolbox^9,32^. The training dataset comprised the top 100 segmentations from the SLIP cohort (restricted to those aged 0-20), based on scores from the automated quality control from SynthSeg+. We consolidated all extracerebral labels, along with unlabeled/non-background voxels, into a single whole-head extracerebral label.

Each BOBs-BCP subject was then registered to the SLIP-Train and CHOP-Path scans, generating augmented synthetic image-label pairs. Each BOBs-BCP subject was registered to each of the 10 SLIP scans using SynthMorph with ANTs GeneralLabel[Linear] transformation^24^. This process generated 710 BCP-SLIP registration pairs. These registered segmentations were fed into the SynthSeg+ brain generator (see Supplement for details) to generate additional synthetic training scans to increase anatomical and contrast variability, yielding 1420 synthetically generated scan and segmentation pairs. The CHOP-path cohort was age-matched by month to the BOBs-BCP individuals, and registration was performed using the same SynthMorph protocol described above, yielding 351 image and segmentation pairs and 702 synthetic pairs.

To increase robustness to differential myelination of the white matter compartment in early brain development, white matter segmentation refinement was performed using K-means clustering (k=2,3,4,5) applied to T1-weighted scans and their corresponding segmentations in BOBs-BCP. This approach simulated different stages of myelination in different white matter tracts by augmenting the white matter segmentation with discrete white matter voxel bundles in the training dataset, to mimic differential myelination of white matter tracts in early brain development. This clustering approach generated 284 white matter clustered segmentations for the BCP dataset, providing more detailed tissue classification within white matter regions. Again, the SynthSeg+ brain generator was used to create an additional 568 synthetic images with corresponding segmentations whereby the newly clustered WM labels were mapped back onto the original left and right white matter labels.

### Training and Testing the ClinSeg Model

Real and synthetic data were combined to create a comprehensive training dataset. The final dataset incorporated the original manual segmentations, registered pseudo-clinical data, refined white matter segmentations, and synthetic augmentations. A 5-fold cross-validation strategy was implemented using nnUNet, with real data reserved for validation while both remaining real and synthetic data were used for training^33^. See the Supplement for a detailed description of the training strategy.

At test time, scans are either kept at their original resolution if already close to isotropic or are otherwise resampled to the target resolution learned by nnUNet (.8x.8x.9) and additionally reoriented to Right, Anterior, Superior (RAS) orientation. We repurposed the existing SynthSeg QC module for automatic quality control, a deep learning model trained to predict the Dice coefficient of segmentations using synthetic data^9^.

### Generating Downstream Cortical Parcellation

We leveraged the FreeSurfer recon-all-clinical pipeline to generate cortical reconstructions based on ClinSeg’s segmentations. The recon-all-clinical pipeline uses a combination of SynthSeg+ for generating segmentations, a deep learning approach for computing distances to the surface (SynthDist), and FreeSurfer functionality for generating cortical parcellations^19,34,35^. We substituted our ClinSeg-derived segmentations in place of the SynthSeg+-derived segmentations. We subsequently refer to this aggregate pipeline as ClinSeg2RAC. In addition to the volumetric segmentations provided by ClinSeg, ClinSeg2RAC provides additional phenotypes (e.g. cortical thickness and surface area) as well as cortical parcellation using any of FreeSurfer’s cortical atlases.

### Evaluating Model Performance Using Datasets With Manual Segmentations

We tested ClinSeg on two independent research-quality datasets with accompanying manual segmentations, Developmental Infant Brain Study (DIBS) and Child and Adolescent NeuroDevelopment Initiative (CANDI), which were not used in model training or validation. DIBS is composed of 47 isotropic T1w scans with segmentations for bilateral cerebral white matter and cortex (0-36 months)^36^. CANDI consists of 101 T1w scans for a pediatric cohort (4-17 years of age)^37^. Segmented regions for CANDI include left and right segmentations for the cerebral cortex, cerebral white matter, cerebellar cortex, cerebellar white matter, lateral ventricles, thalamus, ventral diencephalon (VentralDC), caudate, putamen, pallidum, accumbens, hippocampus and amygdala, brainstem, 3rd and 4th ventricles. For Dice score computation, homologous left and right regions were merged, as were ventricular sub-regions (lateral, 3rd, and 4th), the putamen and pallidum, and the hippocampus and amygdala. Additionally, we compared performance of ClinSeg against BIBSNet 3.4.2 and SynthSeg+^9,16^.

### Evaluating Performance of ClinSeg vs SynthSeg+ in the Scans with Limited Imaging Pathology and Normative Modeling

Using both ClinSeg and SynthSeg+, we generated segmentations for the full CHOP SLIP cohort (n=78,508). Preliminary investigations showed low performance in clinical scans, so we did not assess BIBSNet performance in the full CHOP SLIP sample. ClinSeg segmentations passing the automated SynthSeg QC were then input into the *recon-all-clinical* pipeline to yield cortical parcellations, thickness, and surface area as described above. We modeled global and regional imaging phenotypes using GAMLSS with a Box-Cox t (BCTo) distribution, estimating parameters for mean (µ), scale (σ), skewness (ν), and kurtosis (τ). Fixed effects for µ and σ included sex and log-transformed gestation-adjusted age, modeled using cubic P-splines (pb(), order=3), with smoothing parameters selected via BIC. Age-by-sex interactions were included in the initial model for both µ and σ, and the σ submodel was subsequently simplified using stepwise BIC. Scanner site was included as a random effect for µ and σ. To account for participants contributing multiple sessions, observations were weighted by the inverse of each participant’s session count. The quality of model fit was assessed using visualization of worm plots and quantification of the percentage of data points falling outside the pointwise confidence intervals (see Figure S2)^38^.

### Extending 22q11.2 Deletion Syndrome Case-Control Comparisons With Infant Data

Recent work has leveraged the SLIP sample, along with imaging data from a cohort of individuals with 22q11.2 deletion syndrome (22q11DS) who were scanned at CHOP, in a normative modeling analysis^14^. Specifically, standardized centile *z*-scores for global and regional imaging-derived phenotypes (IDPs) were calculated to estimate case-control effect sizes. However, this analysis utilized the FreeSurfer recon-all-clinical pipeline, which suffers from unreliable segmentation in infants under 6 months of age, precluding use of their data in the analysis. Both to test the stability of the prior findings and overcome the infant data limitation, we replicated the case-control analysis using our ClinSeg pipeline. As in Jung et al, for gray matter volume (GMV) and surface area (SA), a median-based approach was utilized, pooling data across all available acquisition types per subject that passed QC based on SynthSeg composite QC measures and the established SLIP Euler threshold. CT analyses were limited to MPRAGE data (MPR) in individuals aged 1 year or older, prioritizing the single non-contrast MPRAGE scan with the highest Euler score. For subjects scanned at sites not represented in the SLIP cohort, centiles were derived from the fixed-effects component of the fitted penalized-spline GAMLSS models alone; i.e., the site-specific random intercept was set to its expected value of zero.

Linear fixed-effects models were employed to predict each IDP. Diagnosis served as the primary predictor, with adjusted age in days including gestation, sex, and the median Euler number (as a proxy for scan quality) included as covariates. The inclusion of age and sex aimed to address any residual biases not fully accounted for by normative modeling. Following prediction, t-statistics were converted to Robust Effect Size Index (RESI) effect sizes, and all *p*-values were corrected for multiple comparisons using the Benjamini-Hochberg False Discovery Rate (FDR) procedure^21^.

## Data Availability

All data produced in the present study are available upon reasonable request to the authors.

https://openneuro.org/datasets/ds005450/versions/1.0.0

https://www.nitrc.org/projects/candi_share/

## Code and Data Availability

We will be releasing a Docker image and GitHub repository for the ClinSeg tool, along with the growth charts upon publication. GAMLSS analyses and surface plots were performed in R 4.4.0, and subcortex visualizations were generated using the subcortex-visualization toolbox^39^.

## Acknowledgements

This work was funded by NIMH R01MH134896, NIMH R01MH133843, and the CHOP Research Institute. H.F.J.T was funded by NIHR UCLH Biomedical Research Centre Healthcare Engineering and Imaging Theme Early Career Fellowship (ACCESS).

