## Supplemental Material for "ClinSeg: Robust Brain Segmentation for Clinically Acquired Pediatric MRI"

### Supplementary Materials: ClinSeg: Robust Brain Segmentation for Clinically Acquired Infant MRI

**Supplementary Methods** **2**
MedGemma based grading of morphological abnormalities 2
Generating synthetic data using the SynthSeg+ brain generator 2
nnUNetv2 training plans 3
**Supplementary Results** **4**
Milestone comparison across pipelines 11

#### Supplementary Methods

##### MedGemma based grading of morphological abnormalities

Our full set of SQL query search terms included: "ventriculomegaly," "hydrocephalus," "sulcal prominence," "cortical dysplasia," and "encephalomalacia." The MedGemma prompt was as follows:

> *"You are an expert medical AI assistant specializing in radiology. Your task is to analyze the provided report, identify specific neurological pathologies, and classify their severity.*
>
> ***Instructions:***
>
> *1. Carefully read the report provided below.*
>
> *2. Identify whether the following abnormal neurological findings (pathologies) are mentioned - benign enlargement of the subarachnoid space (bess), enlarged extra axial space, sulcal prominence/prominent sulci, chiari malformation, volume loss, ventriculomegaly, encephalomalacia, cortical dysplasia.*
>
> *3. For each finding, assign a severity level from this specific scale: "Mild", "Moderate", "Severe".*
>
> *4. If a condition is mentioned as absent, resolved, or unremarkable, classify it as "Normal".*
>
> *5. Structure your final output as a single, valid JSON object where each key is the pathology and the value is its corresponding severity. Do not include any text before or after the JSON object."*

##### Generating synthetic data using the SynthSeg+ brain generator

For each input segmentation, two synthetic image-label pairs were produced using the SynthSeg+ brain generator.

Parameters for geometric augmentation were chosen as follows: left-right flipping (50% probability), minor scaling (±5%), and small rotations (±5°) were applied, while shearing, translation, and elastic deformation were all disabled. Intensity augmentation included bias field corruption ($\sigma = 0.5$) to simulate scanner inhomogeneity, and resolution randomization to simulate variability across acquisition protocols. Intensity priors were sampled uniformly to maximize contrast diversity. All images were generated at an isotropic output shape of 160 voxels with native resolution preserved.

**nnUNetv2 training plans**

Automated segmentation was performed using the self-configuring nnU-Net v2 framework in its 3D full-resolution configuration. Single-channel images were read with SimpleITK and, based on their foreground intensity distribution, treated as non-CT data and normalized by per-image z-score normalization. Images were resampled to a target spacing of $0.90 \times 0.785 \times 0.785$ mm using third-order spline interpolation (linear for segmentation maps).

The network was a plain convolutional U-Net with 6 stages (32–320 feature channels), each using two $3 \times 3 \times 3$ convolutions with instance normalization and leaky ReLU. Downsampling was isotropic except at the deepest stage, which preserved through-plane resolution. Training used a patch size of $96 \times 160 \times 128$ and a batch size of 2.

Training followed nnU-Net v2 defaults: 1000 epochs per fold with SGD (Nesterov momentum 0.99, initial learning rate 0.01, polynomial decay), a combined Dice and cross-entropy loss with deep supervision, and 5-fold cross-validation. On-the-fly data augmentation (rotation, scaling, Gaussian noise and blur, brightness and contrast, simulated low resolution, and gamma) was applied, with mirroring/flip augmentation disabled.

**Supplementary Results**

| Dataset | Region | ClinSeg dice (mean +- sd) | BIBSNet dice (mean +- sd) | SynthSeg+ dice (mean +- sd) | p-value (Friedman) |
|---|---|---|---|---|---|
| **CANDI** | **Accumbens Area** | **0.5634 ± 0.0649** | **0.6882 ± 0.0637** | **0.6275 ± 0.0718** | **5.730245e-38** |
| **CANDI** | **Brainstem** | **0.8971 ± 0.0116** | **0.9171 ± 0.0185** | **0.9235 ± 0.0130** | **3.069462e-29** |
| **CANDI** | **Cerebellar Cortex** | **0.9333 ± 0.0103** | **0.9557 ± 0.0070** | **0.9327 ± 0.0091** | **1.262170e-33** |
| **CANDI** | **Cerebellar White Matter** | **0.8302 ± 0.0314** | **0.8134 ± 0.0266** | **0.8225 ± 0.0249** | **1.798713e-13** |
| **CANDI** | **Cerebral Cortex** | **0.8813 ± 0.0100** | **0.8676 ± 0.0124** | **0.8482 ± 0.0108** | **2.397607e-38** |

| | | | | | |
|---|---|---|---|---|---|
| **CANDI** | **Cerebral White Matter** | **0.8575 ± 0.0150** | **0.7992 ± 0.0229** | **0.8542 ± 0.0168** | **1.973063e-35** |
| **CANDI** | **Hippocampus+Amygdala** | **0.7926 ± 0.0234** | **0.8140 ± 0.0184** | **0.8367 ± 0.0207** | **4.012131e-38** |
| **CANDI** | **Putamen+Pallidum** | **0.8481 ± 0.0204** | **0.8557 ± 0.0157** | **0.8612 ± 0.0149** | **7.619343e-18** |
| **CANDI** | **Thalamus** | **0.8697 ± 0.0166** | **0.8380 ± 0.0278** | **0.8831 ± 0.0140** | **4.247408e-34** |
| **CANDI** | **Ventral DC** | **0.7744 ± 0.0380** | **0.7851 ± 0.0443** | **0.8144 ± 0.0287** | **1.754542e-21** |
| **CANDI** | **Ventricles** | **0.8072 ± 0.0471** | **0.7682 ± 0.0683** | **0.8116 ± 0.0491** | **9.637595e-30** |
| **DIBS** | **Cerebral Cortex** | **0.9131 ± 0.0269** | **0.8907 ± 0.0363** | **0.8595 ± 0.0140** | **4.218169e-17** |
| **DIBS** | **Cerebral White Matter** | **0.9085 ± 0.0267** | **0.8896 ± 0.0367** | **0.8574 ± 0.0132** | **1.227522e-16** |

**Table S1. Dice scores across segmentation models and regions for datasets with manual segmentations.**

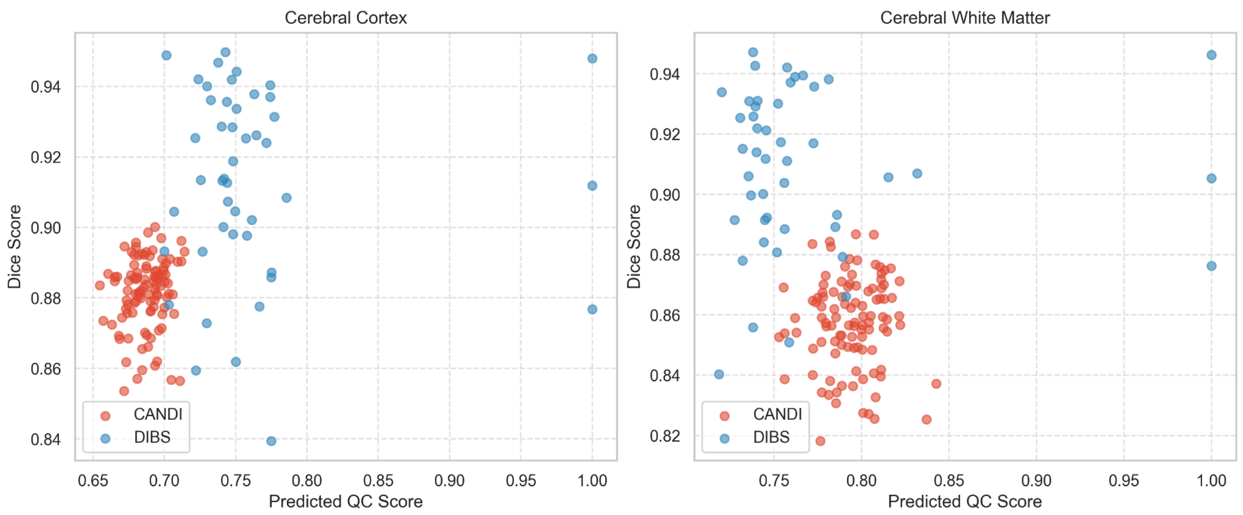


**Fig S1. Concordance between Dice scores and predicted QC scores for ClinSeg segmentations versus manual segmentations in the DIBS and CANDI datasets.** The cut-off for a passing QC score is 0.65 in each region.

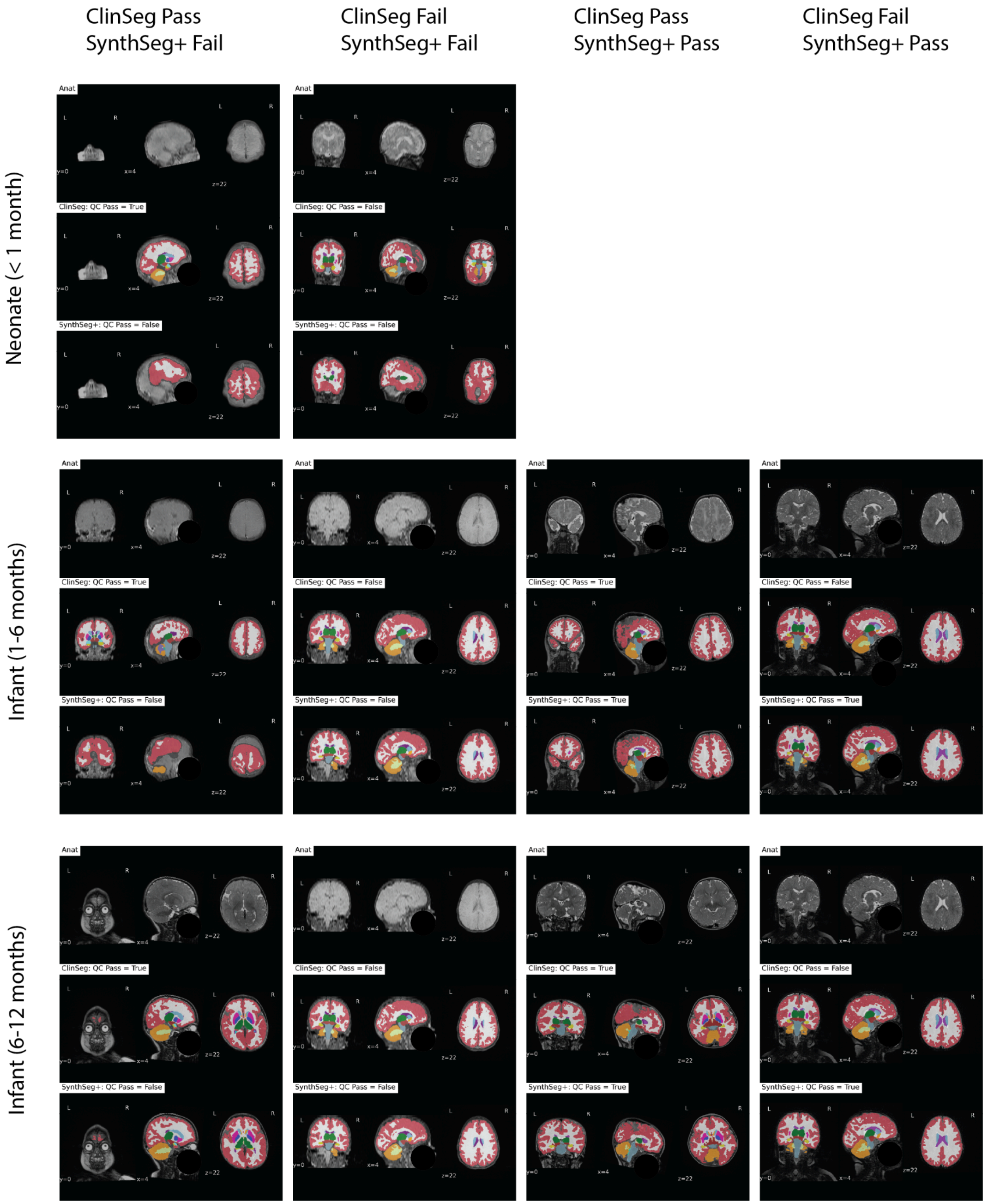


**Figure S2. ClinSeg vs SynthSeg+ segmentations in SLIP.** We show example scans and accompanying ClinSeg vs SynthSeg+ segmentations across three age bins (neonates, infants between 1-6 months of age, and infants between 6-12 months of age).

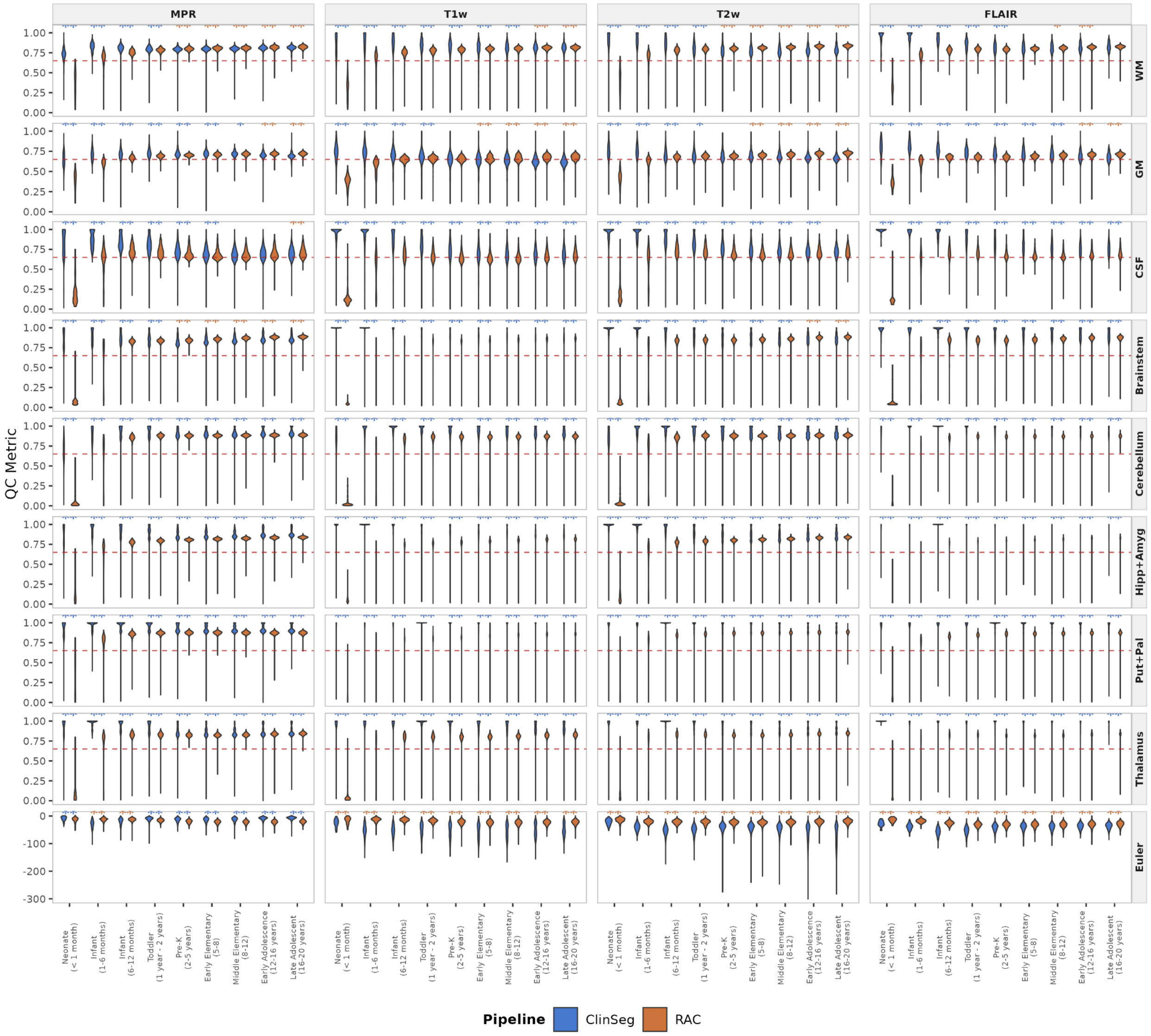


**Figure S3**. **Segmentation and surface reconstruction quality control measures across age categories and acquisition types.**

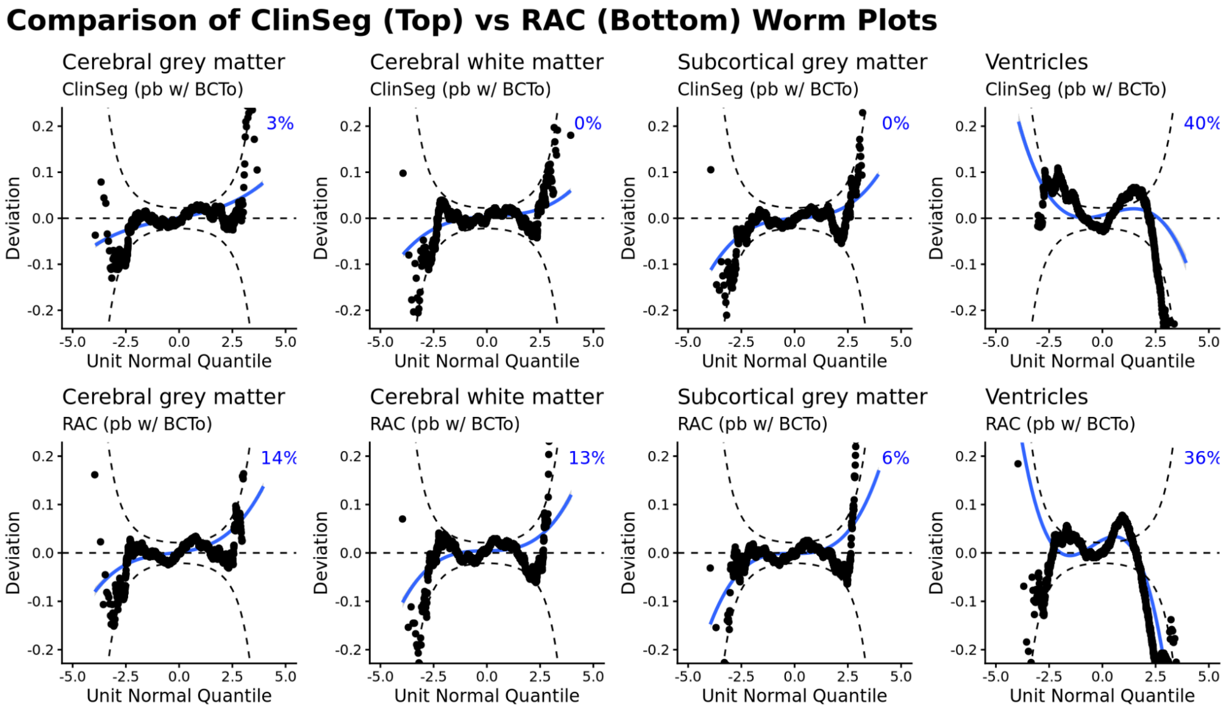


**Figure S4**. **Wormplots across composite segmentation structures for ClinSeg (top) versus RAC (bottom) GAMLSS models fit using penalized splines.** A wormplot is a detrended normal Q-Q plot used to visually diagnose the fit of GAMLSS models. These examples of wormplots show  the penalized spline models fit using the BCTo distribution for global measurements following cortical refinement. Percentage in blue refers to the percentage of data points outside of the plotting window.

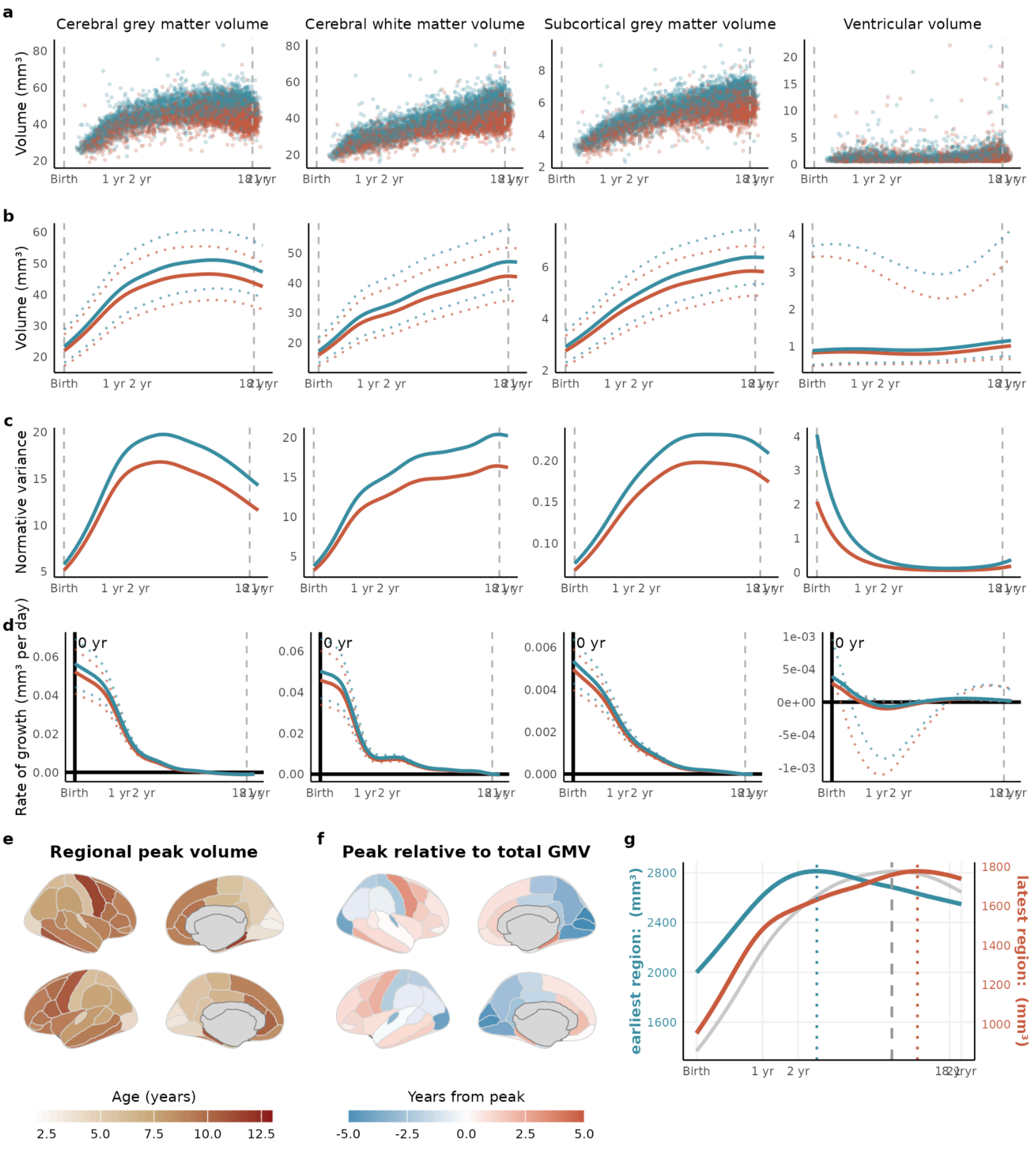


**Figure S5. Clinical growth charts constructed using RAC derivatives with penalised splines & BCTo family. (a)** Raw bilateral cerebrum tissue volumes for grey matter, white matter, subcortical grey matter and ventricles are plotted for each session's data as a function of age (log-scaled) with points colored by sex. **(b)** Normative brain-volume trajectories were estimated using GAMLSS modeling as described in the Methods, with trajectories stratified by sex (female, red; male, blue). The dotted trajectories represent the 2.5% and 97.5% centiles. **(c)** Trajectories of median between-subject variability are estimated as the approximated predicted dispersion. **(d)** Rates of volumetric change for each sex-specific trajectory was estimated using the first derivative of the median volumetric trajectory, as described in Bethlehem et al. Solid horizontal lines denote the tissue-specific volume at which the tissue begins to shrink while the solid vertical line denotes the age of maximum growth. **(e)** Regional peak volumes for the 68 Desikan-Killiay cortical regions. **(f)** Peak relative to GMV peak, where GMV peak is subtracted from each regional peak. **(g)** Trajectories for the earliest peaking region (right pericalcarine, blue line) and the latest peaking region (right parahippocampal, red line), showing the range of regional variability relative to the GMV trajectory (grey line). Regional volume peaks are denoted as dotted vertical lines either side of the global peak, denoted as a dashed vertical line, in the bottom panel. The left *y* axis on the bottom panel refers to the earliest peak (blue line); the right *y* axis refers to the latest peak (red line).

**Milestone comparison across pipelines**

Comparative analysis across the non-linear modeling frameworks and segmentation pipelines revealed distinct tissue-specific peaks. Using p-splines, the ClinSeg-derived GMV peaked later (8.95 years) than the RAC-derived model (8.09 years). Of note, under the fractional polynomial framework, the ClinSeg2RAC pipeline shifted the GMV peak earlier to 6.9 years, aligning more closely with the 5.9-year GMV peak reported for the LBCC (Figure S4). WMV showed more divergent results compared with Bethlehem et al, with ClinSeg p-spline models revealing a more adolescent peak of 18.1 years (RAC: 16.7 years) compared with the peak of 28 years reported previously. Subcortical gray matter volume (sGMV) peaked in adolescence in the LBCC sample (14.  years), and RAC recapitulated this peak more closely at 17.10 years while ClinSeg had a later peak of 20.9 years. Finally, accurate modeling of ventricular volume is contingent on a lifespan sample which would capture the exponential ventricular expansion documented by the LBCC, and both ClinSeg and RAC hit premature peaks of 20.9 years. Compared with the LBCC reported age of 2.4 years for WMV maximum rate of growth, we found a substantially earlier result of 0.1 years for ClinSeg (0.2 years for RAC), potentially reflecting the younger nature of our dataset and more early life data available.

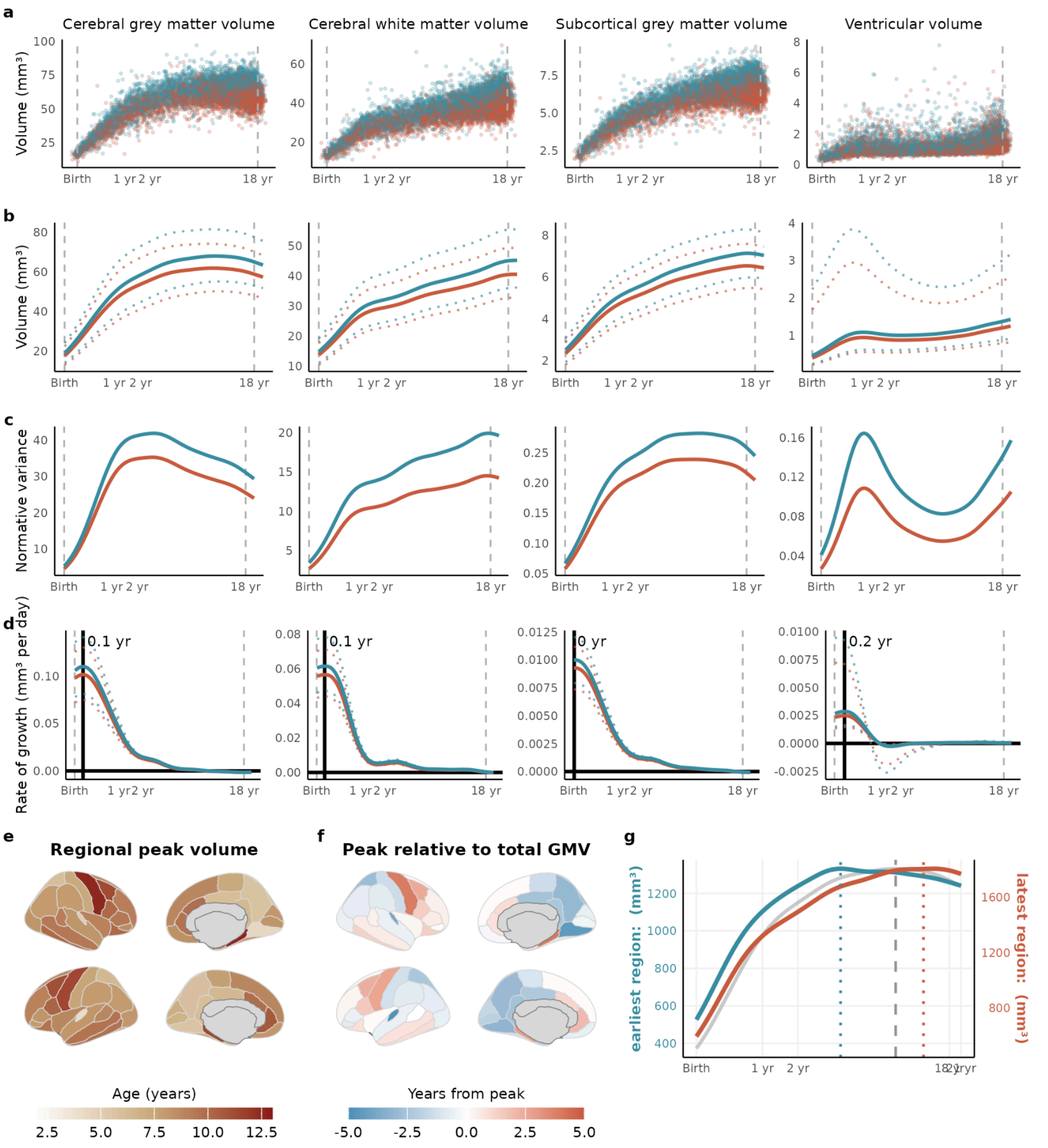


**Figure S6. Clinical growth charts for composite regions constructed using ClinSeg derivatives (penalized splines) before RAC cortical refinement. (a)** Raw bilateral cerebrum tissue volumes for grey matter, white matter, subcortical grey matter and ventricles are plotted for each session's data as a function of age (log-scaled) with points colored by sex. **(b)** Normative brain-volume trajectories were estimated using GAMLSS modeling as described in the Methods, with trajectories stratified by sex (female, red; male, blue). The dotted trajectories represent the 2.5% and 97.5% centiles. **(c)** Trajectories of median between-subject variability are estimated as the approximated predicted dispersion. **(d)** Rates of volumetric change for each sex-specific trajectory was estimated using the first derivative of the median volumetric trajectory, as described in Bethlehem et al. Solid horizontal lines denote the tissue-specific volume at which the tissue begins to shrink while the solid vertical line denotes the age of maximum growth. **(e)** Regional peak volumes for the 68 Desikan-Killiay cortical regions. **(f)** Peak relative to GMV peak, where GMV peak is subtracted from each regional peak. **(g)** Trajectories for the earliest peaking region (left transverse temporal, blue line) and the latest peaking region (right parahippocampal, red line), showing the range of regional variability relative to the GMV trajectory (grey line). Regional volume peaks are denoted as dotted vertical lines either side of the global peak, denoted as a dashed vertical line, in the bottom panel. The left *y* axis on the bottom panel refers to the earliest peak (blue line); the right *y* axis refers to the latest peak (red line).

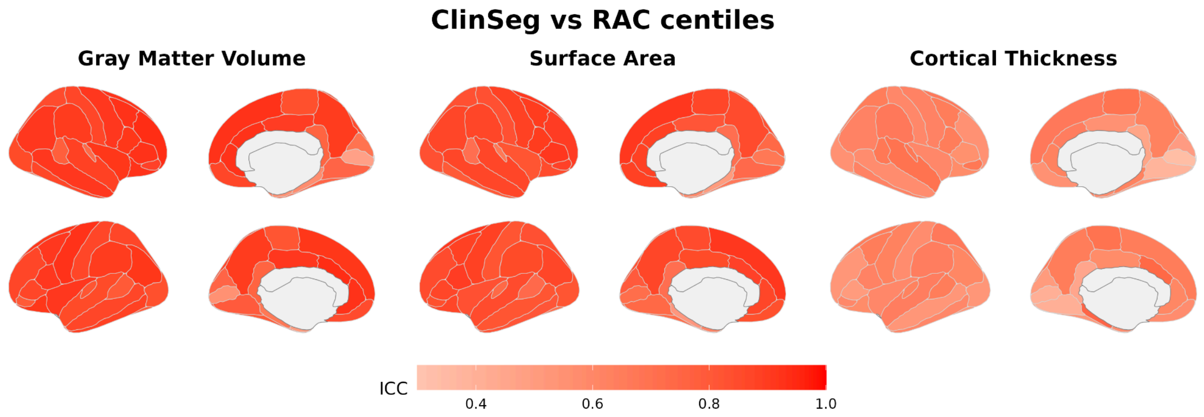


**Figure S7. Regional ICC for ClinSeg vs RAC centiles in the SLIP dataset.** For subjects who survive QC for RAC and ClinSeg, we compared their centiles using ICC. Median models used for the grey matter volume and surface area comparison while the MPR model is used for the cortical thickness comparison.

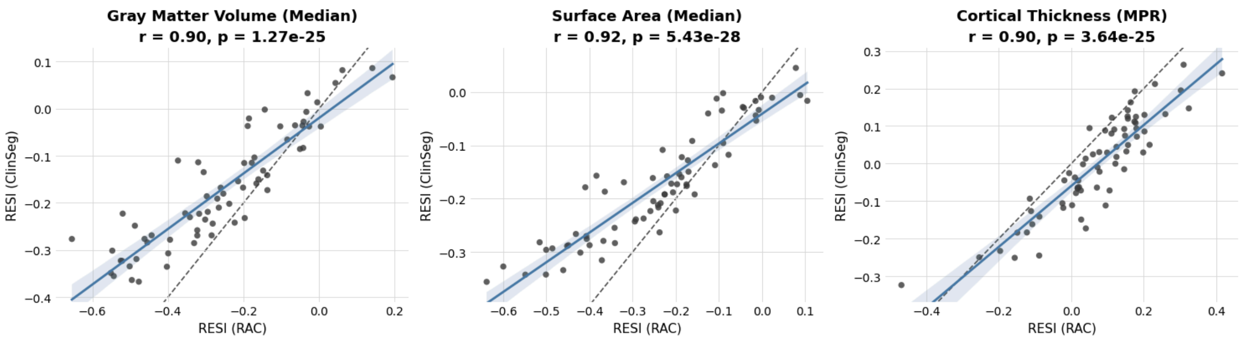


**Figure S8. 22q11DS effect size comparison across Jung et al 2026 and ClinSeg based results.**

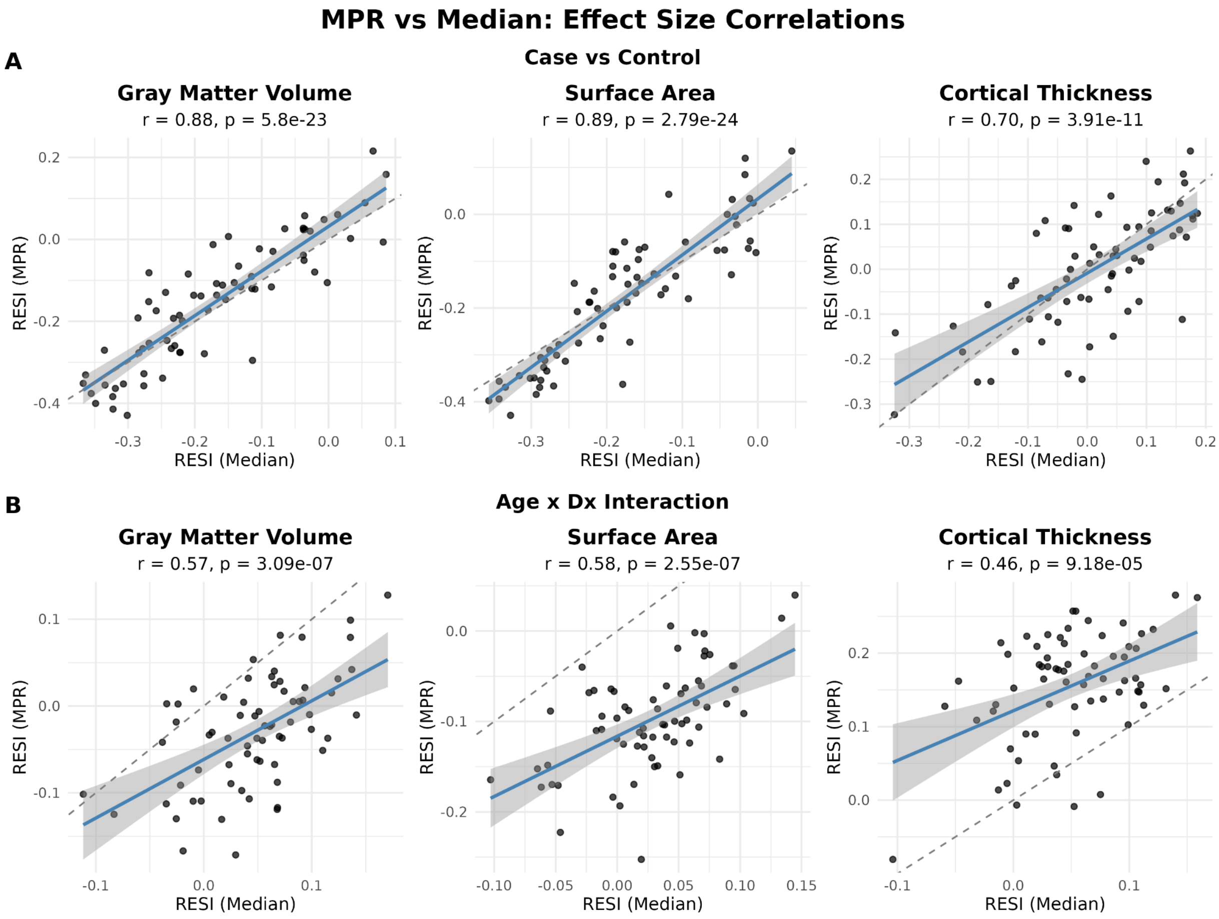


**Figure S9. 22q11DS effect size comparison across case-control and age by diagnosis interaction analyses.**

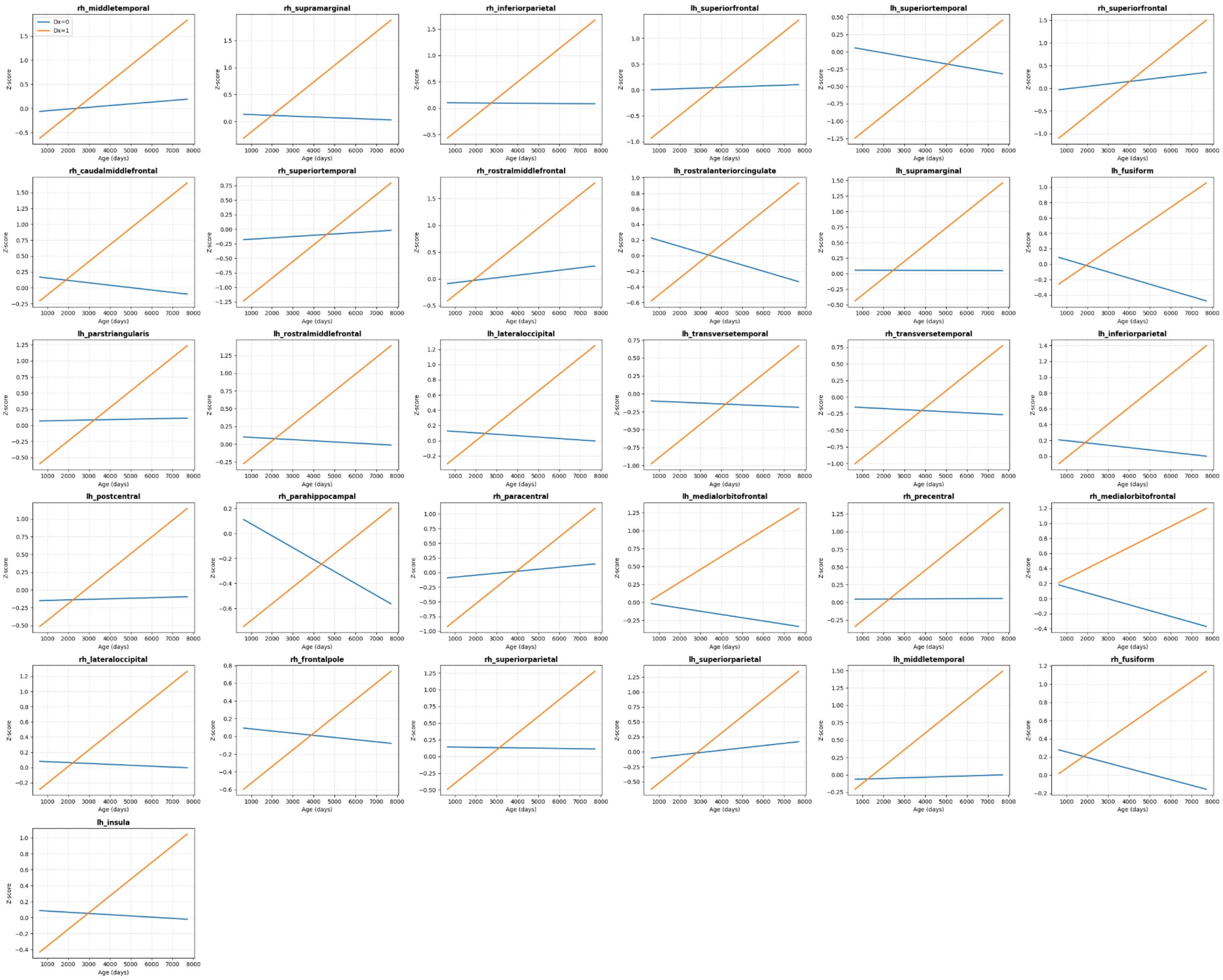


**Figure S10. Age by 22q11DS diagnosis interaction plots.** Ordinary least squares (OLS) regression trajectory plots for controls (Dx 0, blue) vs individuals with 22q11DS (Dx 1, orange) for normalized cortical thickness centile z scores. Covariates (sex and Euler number) are held constant for visualization purposes. While the control trajectories are static or slightly declining in most cases, the 22q11DS predicted trajectories have a more positive slope.
